# A Decision Analytic Approach for Social Distancing Policies During COVID-19 Pandemic

**DOI:** 10.1101/2020.06.24.20139329

**Authors:** Zeynep Ertem, Ozgur M. Araz, Mayteé Cruz-Aponte

## Abstract

The COVID-19 pandemic has become a crucial public health issue in many countries including the United States. In the absence of the right vaccine strain and sufficient antiviral stockpiles on hand, non-pharmaceutical interventions have become valuable public health tools at the early stages of the pandemic and they are employed by many countries across the globe. These interventions are designed to increase social distancing between individuals to reduce the transmission of the virus and eventually dampen the burden on the healthcare system. The virus transmissibility is a function of the average number of contacts individuals have in their communities and it is highly dependent on population density and daily mobility patterns, along with other social factors. These show significant variation across the United States. In this article, we study the effectiveness of social distancing measures in communities with different population density. Specifically, first we show how the empirical estimation of reproduction number differs for two completely different states, thus the experience of the COVID-19 outbreak is drastically different, suggesting different outbreak growth rates in practice. Second, we develop an age-structured compartmental model for simulating the disease spread in order to demonstrate the variation in the observed outbreak characteristics. We find that early trigger and late trigger options present a trade-off between the peak magnitude and the overall death toll of the outbreak which may also vary across different populations.

## 1 Introduction

On March 11, 2020, the World Health Organization (WHO) declared the global outbreak caused by Coronavirus Disease 2019 (COVID-19) as a pandemic. This new virus is of the coronavirus family, named SARS-CoV-2 (CDC 2020b) which was first detected in Wuhan, China, on December 2019, and quickly spread around the world. Currently, the total number of people diagnosed with COVID-19 has exceeded six million and it is present in nearly every country. As of June 2020, this number continues to rise with corresponding fatalities (CDC 2020c). Although there are still several unknowns regarding the natural characteristics of the virus, the main mode of transmission is identified to be mainly through respiratory droplets expelled from the mouth or nose of an infected person and subsequently inhaled by a susceptible person. It is also possible to get COVID-19 through contact with contaminated surfaces as well (CDC 2020a). Such modes of transmission creates a disease dynamic that is easy to grow. In addition, early epidemiological estimates show that the severity of this novel virus is relatively very high in elderly population than younger populations (Verity *et al*. 2020).

The COVID-19 pandemic has become a crucial public health issue in many countries including US. In the absence of the right vaccine strain and sufficient antiviral stock piles on hand, non-pharmaceutical interventions have become valuable public health tools at the early stages of the pandemic and employed by many countries across the globe including the US. These interventions are designed to increase social distancing between individuals to reduce the transmissibility and eventually dampen the burden on the healthcare system so that the overall risk of public health is minimized. The effectiveness of these social distancing measures depends on the population density and underlying mobility patterns of a region. In the United States (US), the states show significant variation in population density and demographic configurations. For example, when Nebraska is compared to New York the population density and the mobility patterns are drastically different. Nebraska has low population density and mobility patterns with low-use of public transportation. New York, on the other had, especially New York City exhibit high population density and high-use of public transportation.

In this article, we study the practical effectiveness of both social distancing measures for outbreak in regions with different population density and mobility patterns as well as reopening strategies. Specifically, first we show that the evolution of the empirical fit for reproduction number (i.e. a measure for transmissibility) for two different states in the US, i.e., Nebraska and New York, for the COVID-19 outbreak are drastically different, suggesting different outbreak transmission rates in practice, emulating different states. Second, we develop an age-structured compartmental model to simulate the disease spread for different growth rates and show that the observed outbreak characteristics are different for these states. Third, employing computational experiments with the simulation model we show that social distancing measures with different decision parameters and paths (e.g., various triggers to start and different lengths of social distancing) can result in effective management of the outbreak in dissimilar states. We find that early trigger and late trigger options present a trade-off between the peak magnitude and the overall death toll of the outbreak. Finally, we analyze reopening strategies with a two-phased reopening scenario. We show that for outbreaks with small transmission rate a short length of social distancing and immediate reopening scenario may be feasible. Whereas, for outbreaks with medium or high transmission rate the longer the phases until the re-opening the more dampened the burden of the outbreak; and the further the timing of the peak, which may provide more time for the public health officials to increase healthcare capacity.

## 2 Literature Review

While the early epidemiological estimates about the natural progression of the disease were reported with significant uncertainty, the public health authorities and researchers around the globe reported their surveillance data and findings that provided details about case fatality rates and transmissibility. An early study on the transmission dynamics of COVID-19 analyzed data of the first 425 confirmed cases in Wuhan, China and found that the mean incubation period was 5.2 days. However, the 95^th^ percentile of the incubation period was 12.5 days in this sample (Li *et al*. 2020a). In addition, this study estimated the basic reproductive number (*R*_0_), which is the average number of secondary cases generated from a single infectious case in a completely susceptible population, as anywhere between 1.5 and 3.5 (Li *et al*. 2020a). These epidemiological estimates show that the virus strain is highly transmissible and can infect mass populations across the globe in a very short amount of time. Another study on news reports and press releases about COVID-19 outside Wuhan estimated the mean incubation period and 97.5^th^ percentile as 5.1 days and 11.5 days, respectively (Lauer *et al*. 2020). Linton *et al*. (2020) also estimated the expected incubation time around 5 days with relatively higher range of 2 to 14 days. Given relatively high estimates of the basic reproductive number *R*_0_, and relatively long incubation period, there is an exponential growth of confirmed COVID-19 cases in several countries and developing control measure for the epidemic is quite challenging. More importantly, a significant proportion of cases are reported to be asymptomatic but infectious cases (Li *et al*. 2020a). This characteristic of the virus is found one of the main drivers of the spread of COVID-19 across populations. Therefore, social distancing measures are crucial for gaining time for healthcare systems to meet the demand for care and ultimately mitigating the impact of the pandemic.

By mid-March, there has been a significant impact on the economy of several countries as more restrictions to mitigate the risk are being implemented, such as travel bans, cancellation of social events (concerts, sports, etc.), closure of non-essential businesses, “stay at home” orders. While social distancing interventions have been implemented in various states of the US and also in other countries worldwide they have been very controversial policy since they can have significant impact on the economy. Because it is usually hard to accurately estimate the transmissibility and severity of infections caused by a newly emerged virus at the early stages of the epidemic, there will still be many uncertainties about disease progression dynamics in the communities. Therefore, the public health decision makers will have to make important decisions on using school closures and other social distancing measures as community mitiga-tion strategies until the right strain of a vaccine is developed and distributed. However, these strategies may vary based on the social and demographic characteristics of different communities. In this article, we present the timely evolution of the estimates of the reproductive number in two different states, i.e., New York where we observe high transmission rates; and Nebraska where we observe relatively lower community transmission rates, along with a baseline scenario based on more global estimates. We evaluate social distancing measures to analyze whether one unified policy would be effective in completely different states in the United States.

Social distancing measures have significant impact on the spread of infectious diseases in populations, this is also observed during the COVID-19 pandemic (Courtemanche *et al*. 2020). Recently, in the literature there are several analytical approaches developed to analyze effectiveness and/or cost effectiveness of various public health mitigation strategies. Fumanelli *et al*. (2016) consider school closure based social distancing interventions in which closure strategies are based on school absenteeism: nationwide, countywide, reactive school-by-school and reactive gradual. Gojovic *et al*. (2009) developed a simulation model for H1N1 2009 outbreak in a structured population in Ontario and evaluated different mitigation strategies. In their analysis the decision analytic framework is used for different mitigation strategies including school closures, however, these analyses are limited and do not involve any cost effectiveness analysis. Ciavarella *et al*. (2016) studied school closure policies at municipality level for mitigating influenza spread using compartmental models. Decision analytics and support systems are used with compartmental model to control infectious disease epidemics.

While a pandemic possibility continuously posed global risks to public health systems and business continuity (Araz *et al*. 2020), federal and state health departments developed public health policies for mitigation and response (Ramirez-Nafarrate *et al*. 2019). While modeling and simulation studies are used for optimal pharmaceutical intervention design, which include vaccination policies (Duijzer *et al*. 2018b,a), non-pharmaceutical interventions are also modeled with disease progression dynamics (Griffiths *et al*. 2000, Teytelman & Larson 2012). In developing social distancing policies, researchers and public health officials have used models and quantitative analyses to evaluate their effectiveness and costs under possible pandemic scenarios. Some studies demonstrate school closures can have a significant impact on the effective reproduction number and on the overall spread of disease (Lee *et al*. 2010). One advocates, for example, 26 weeks of school closure in conjunction with other policies (Sander *et al*. 2010). Since the H1N1 influenza pandemic in 2009 many studies showed the cost effectiveness and epidemiological impact of a large set of school closure policies. Different than the H1N1 pandemic in 2009, COVID-19 pandemic has raised more political, social and economical challenges as it turned to be a more severe pandemic. Therefore, in this article we evaluate several more comprehensive social distancing policies coupled with different strategies to reopen. The outcomes measures considered for the policies are health care utilization and fatalities via a model that is calibrated based on state specific transmission data and socio-demographic decomposition. This study contributes to the literature by presenting a data driven epidemiological modeling with decision analytics framework to inform social distancing policies for different states. In addition the analysis show that multiple waves of infections should be expected based on the social distancing policies, which would vary in peak magnitude and timing depending on the policies. Finally, the study highlights how time gained by using social distancing interventions can vary across states with different population configurations until mass vaccination or antiviral dispensing can become available.

## 3 Model

In this paper, we present a decision analytic approach for social distancing during a pandemic. We develop and simulate an age structured mass action model integrated with a decision analytical approach to evaluate the impact of social distancing measures and reopening strategies. The impact of these strategies are evaluated based on the computational results obtained for cumulative attack rate, projected peak value of infections, i.e., peak prevalence (%), timing of the peak, peak hospitalizations, and cumulative deaths.

In our decision analytic approach, the social distancing measures are triggered with the prevalence in the community similar to Araz *et al*. (2012), as most of the social distancing policies are implemented by monitoring the % of cases as triggers. All the considered reopening strategies that are evaluated are modeled after a fixed duration of social distancing policies and with a gradual removal strategy implemented based on multiple phases. We assume the social distancing policies are implemented based on the prevalence of COVID-19 infections in the community and a series of reopening decisions are considered after a fix duration of social distancing. The considered closure durations are 1, 2, 3, 4, 8, 16 and 24 weeks which are coupled with the prevalence based triggers, i.e., 0.001%, 0.05%, 0.1%, 0.2%, 0.3%, 0.4%, 0.5% and 1%. See Figure 2 for all the scenarios considered in our analyses. The removal of the policies are modeled in phases, following the guidelines released from CDC (2020b). Next, we first present the differential equation based disease spread model constructed to simulate disease progression in the communities, then we explain the triggering strategies used for social distancing policies and phase-based reopening.

### 3.1 Disease Spread Model

Compartmental disease spread models are widely utilized in computational and mathematical epidemiology (Anderson & May 1991). In these models all individuals act similarly but separately from each other in a homogeneously mixed population (Dimitrov *et al*. 2009). Here, in this study we use an age-structured, continuous time compartmental model using a population specific data and considering uncertainty on several input parameters. The equations (1)-(7) model the transmission dynamics of COVID-19 disease in a given population and flow of individuals moving from one disease state to another as given in Figure 1. They represent the disease progression for individuals who are first susceptible then exposed to the disease, and then become asymptomatic or symptomatically infectious. Asymptomatic infectious individuals either develop symptoms and become symptomatically infectious or recover from the disease without any symptoms. The symptomatic infectious individuals can either recover themselves or be hospitalized. The hospitalized individuals can either recover or die from the disease. *S*_*i*_(*t*) represents the number of susceptible individuals in the community at time *t* and for age group *i, E*_*i*_(*t*) represents the exposed individuals, while *I*_*A*_ (*t*) is used for asymptomatic infectious individual and *I*_*S*_ (*t*) is used for the symptomatic infectious cases. *R*_*i*_(*t*) is used for the recovered cases in age group *i* and *H*_*i*_(*t*) is the hospitalized cases at time *t* for the age group *i*. Finally *D*_*i*_(*t*) represents the number of deaths in each age group at time *t*. The force of infection for each age group is represented with *λ*_*i*_, and it is computed as 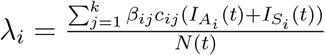.

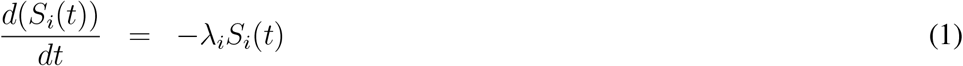

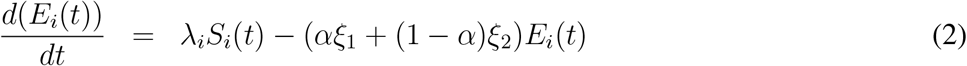

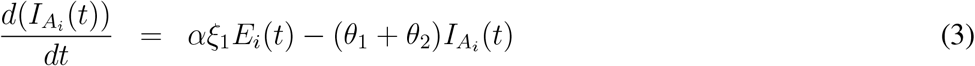

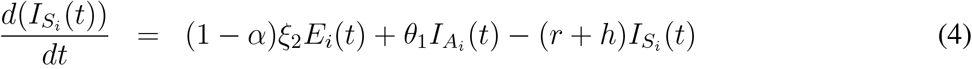

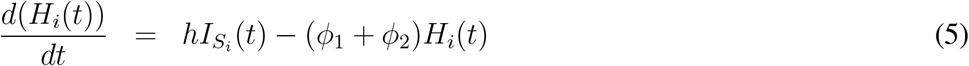

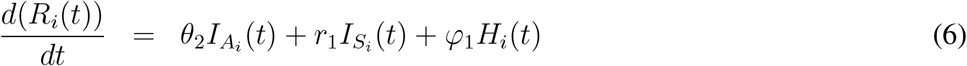

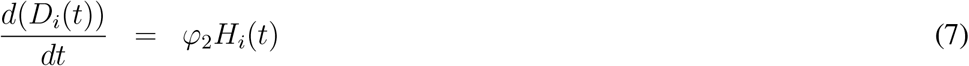

**Figure 1:**
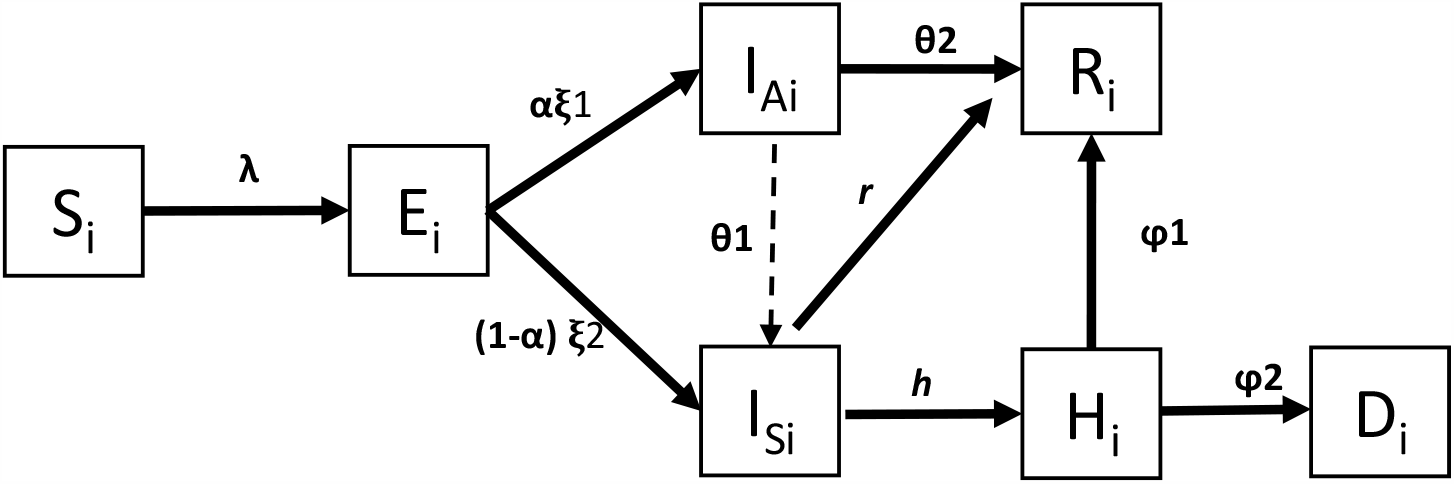
Natural progression of COVID-19 with hospitalization

Using the model presented above we have derived the basic reproductive number *R*_0_ of the system, which is age specific, since the model is an age structured one. Using the next generation operator the theoretical expression for *R*_0_ is derived as given below. See the Appendix B for the details of the derivation process.

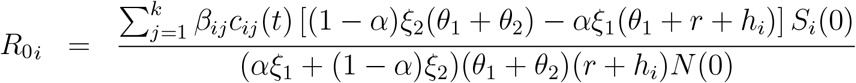

. Now, the time dependent reproductive number i.e., *R_i_*(*t*) for age group i can be stated as following:

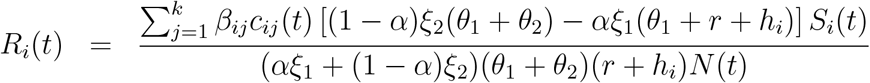

**Proposition 1** *The epidemic control can be achieved if R*_*i*_(*t*) *≤* 1 *can be satisfied with social distancing policies for all age groups*.

We calibrate the model based on the daily cases reported for considered communities to achieve the observed disease propagation and using the result stated in proposition 1 we derive community specific analysis for evaluating the social distancing policies.

### 3.2 Modeling Closure and Reopening Decisions

We evaluate a range of social distancing and reopening strategies for different scenarios for the transmissibility and estimated ranges of severity of the pandemic. Each social distancing policy alternative consists of a prevalence-based trigger and a fixed duration until a reopening decision is made as shown in Figure 2.

**Figure 2:**
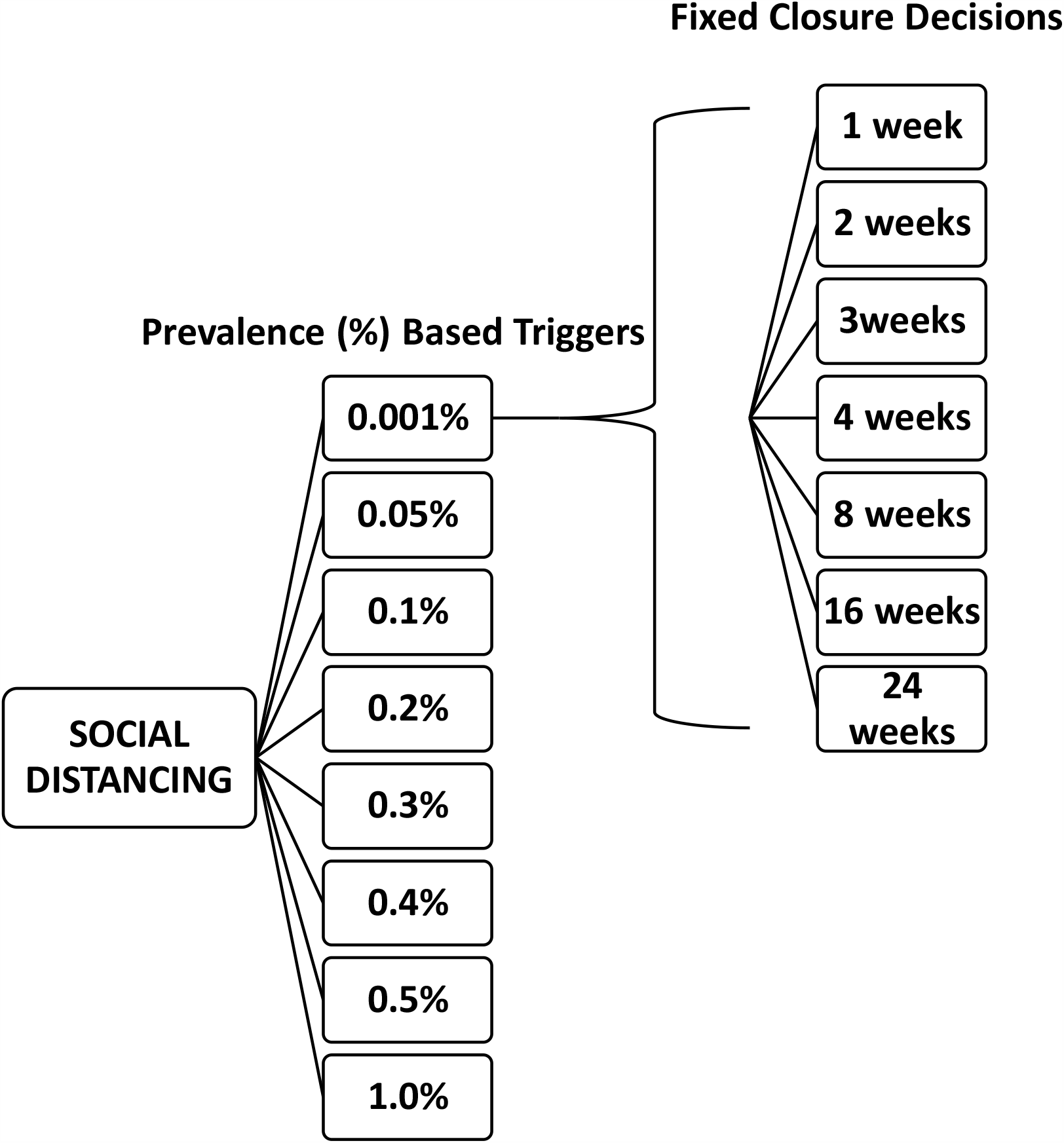
Social Distancing Triggers and Duration Options

Now we explain the modeling process of closure and reopening decisions. As defined earlier 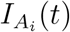 represents the asymptomatic infections for age group *i* at time *t* and *I*_*S*_ (*t*) represents the symptomatic infections for age group *i* at time *t*. In our model, age groups *i* ∈ {*k, a, e*} are defined as *k* for ”kids”, *a* for ”adults” and *e* for ”elderly”. Let the function *f* (*t*) be the cumulative number of individuals infected at time *t* which is computed as following:

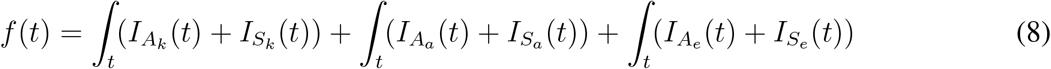

We can mathematically define trigger time for social distancing policy to take place as follows:

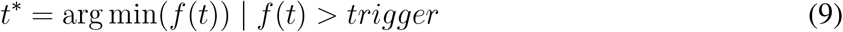

Given the policy trigger time, *t\**, below we show how contact rates *c*_*ij*_(*t*) for each age group changes at time *t* as the closure and reopening decisions are implemented. As the effects of policies would take some time to be observed, let *δ* be the implementation time after the trigger is hit.

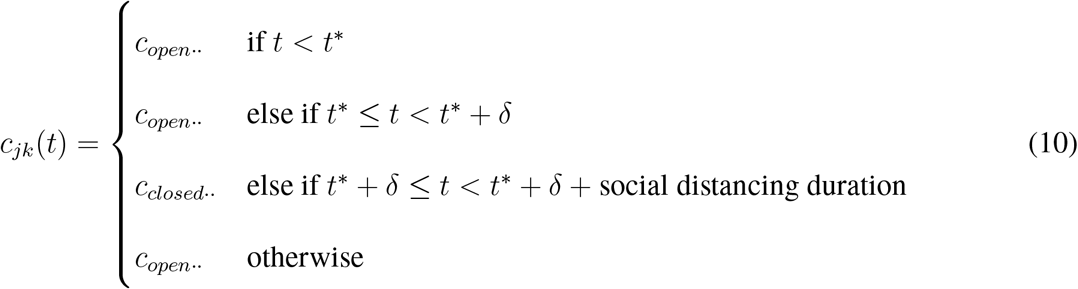

### 3.3 Data

The early published estimates of the basic reproduction number (*R*_0_) of the COVID-19 were slightly higher than 2 (Li *et al*. 2020a). However, depending on the source of the data used in these estimation studies, there were some variation in these estimates. Beside the basic reproductive number, other estimates of the key parameters used in the model are presented in Table 1 which are compiled from the literature and include the latency period, proportion of the asymptotically infected individuals, age dependent hospitalization and mortality rates. Using the cumulative cases data from each state we considered in this study, i.e., New York and Nebraska we have estimated respective reproduction numbers. Then, using the theoretical formula for *R*(*t*) we have calibrated contact rates to achieve the observed reproduction of the cases estimated for each state. See Appendix Table 8 for the values used for the model calibration.

**Table 1:** Model Parameters

| Definition of Parameters | Notation | Value | Range | References |
| --- | --- | --- | --- | --- |
| Force of infection | *computed* | - | - | - |
| Basic reproductive number | $R_0$ | 2.1 | [1.5, 3.15] | - |
| Transmission between age groups | $\beta_{ij}$ | A. Table 7 | — | - |
| Age specific contact rates | $c_{ij}$ | A. Table 8 | — | Mossong *et al.* (2008) |
| Asymptomatic latency period | $\xi_1$ | 5 days | — | Li *et al.* (2020b) |
| Symptomatic latency period | $\xi_2$ | 5 days | — | Li *et al.* (2020b) |
| Asymptomatic proportion | $\alpha$ | Uniformly distributed | 10 – 34.8% | CDC, 2020<br>Mizumoto *et al.* (2020) |
| Rate of asymptomatic developing symptoms | $\theta_1$ | 5% | — | Li *et al.* (2020b) |
| Asymptomatic recovery period | $\theta_2$ | 7 days | — | Li *et al.* (2020b) |
| Symptomatic recovery period | $r$ | 7 days | — | Li *et al.* (2020b) |
| Age specific hospitalization rate | $h_i$ | Uniformly distributed | A. Table 9 | CDC, 2020 |
| Age specific recovery rate | $\varphi_1$ | A. Table 9 | — | CDC, 2020 |
| Age specific mortality rate | $\varphi_2=1-\varphi_1$ | A. Table 9 | — | CDC, 2020 |

In this article, we use daily number of cases data published by the CDC from the January 3rd, 2020 to April 23rd, 2020 for the states of New York and Nebraska and calibrated the contact rate parameter value to achieve the reproductive number estimated for each state. The cumulative number of case over time for each state are presented in Figure 3, with respective time dependent reproduction number estimates.

**Figure 3:**
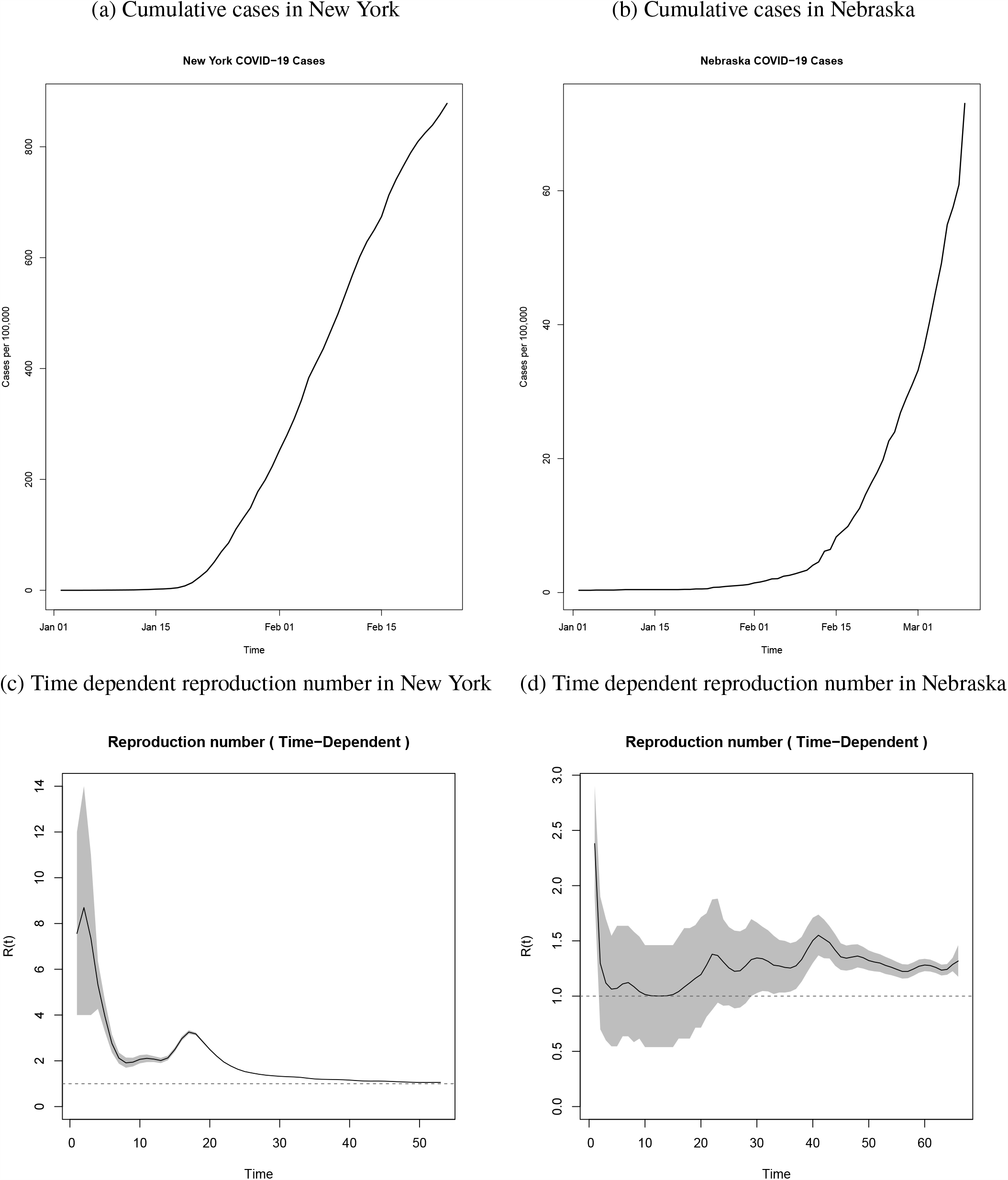
Cumulative Cases in New York and Nebraska by the end of April, 2020

## 4 Results

Using our compartmental model, we evaluate different social distancing strategies by varying two key parameters, namely the threshold prevalence to start the social distancing policies and the length of social distancing. We also evaluate these strategies for different *R*_0_ values (i.e., epidemic growth rate), as empirically we find different growth rate values for different states, in addition to a base case of globally estimated *R*_0_ value of 2.1.

For example, the case reproduction dynamics in New York is different than the one in Nebraska. New York has higher transmission rate and Nebraska has lower transmission rate than the base scenario (*R*_0_ = 2.1). Figure 4a shows the temporal dynamics of outbreak for the three different growth rates with *R*_0_ values of 1.57, 2.1 and 3.15, when no social distancing policy is executed. i.e., labeled as ”no intervention”.

**Figure 4:**
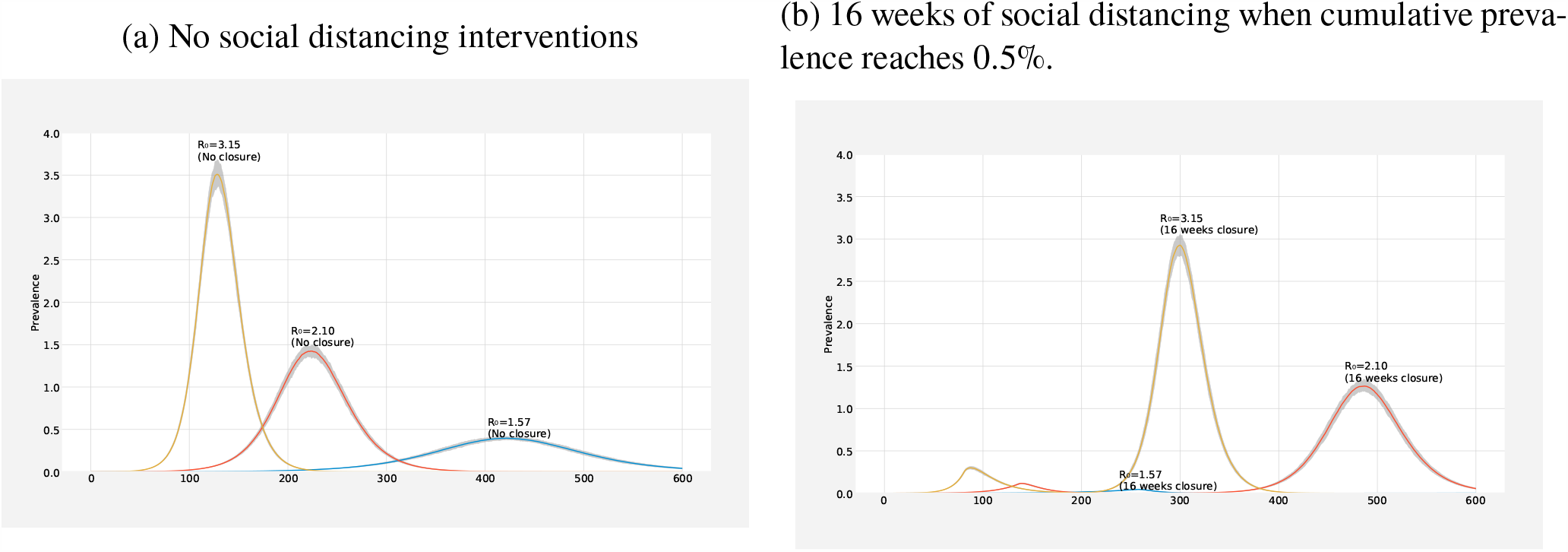
The graph compares different *R*_0_ values, 1.57, 2.1 and 3.15. Lines indicate the median value and the shading indicates the inner 95% range of values of the 100 simulations. Peak timing and magnitude of the pandemic depends on the *R*_0_ values.

We find that practical growth rate of the outbreak in a region directly affects the dynamics of the outbreak. Regions with high transmission rate should expect to experience their peak to be observed much earlier than regions with lower transmission rate. Similarly, the magnitude (i.e. peak height) of the outbreak is significantly higher for those regions with high growth rate than those with small growth rate, as expected.

Furthermore, when same social distancing policies are employed for these different states, we observed different effects of the interventions. For example, Figure 4b compares one specific social distancing policy, i.e., triggering when the cumulative outbreak reaches 0.5% infections under different basic reproductive number scenarios. For states with higher growth rate, the outbreak results in higher and earlier first peak, and an earlier second peak. For regions with lower reproduction number, the social distancing trigger threshold is reached later and the observed peak would be smaller. These simulations suggest that the same social distancing policy can have different effects on the evolution of the epidemic in different populations. Thus, we evaluate each state separately. In the following sections, we evaluate base scenario with (*R*_0_ = 2.1), high transmission rate scenario with (*R*_0_ = 3.15) and low transmission scenario with (*R*_0_ = 1.57) in order to reflect the time dependent reproduction number estimates using COVID-19 cumulative cases data from each state.

### 4.1 Base Scenario (*R*_0_ = 2.1)

Early estimates for the characteristics of the COVID-19 virus suggest that basic reproductive number would be greater than 2, however wide uncertainty rages are also reported (Li *et al*. 2020b). Therefore we use a base case scenario for the transmission dynamics with an *R*_0_ value of 2.1. One critical question in mitigation efforts is to decide when to start the social distancing. Figure 5 shows how applying two distinct trigger thresholds, i.e., 0.5% and 1%, can change the dynamics of the outbreak with a fixed duration of social distancing under the base case scenario with 8 weeks of social distancing. Specifically, the graph compares early trigger (0.5%) and late trigger (1%) options with no intervention. When the social distancing policy is triggered early (0.5%), the outbreak has a smaller first peak in number of cases, and a much higher second peak. Conversely, when the social distancing policy is triggered late (1%), the outbreak has a higher first peak, and a smaller second peak. The early trigger results in an earlier second peak than when a late trigger is used. In both cases, the peaks are considerably dampened compared to the peak without any social distancing implemented.

**Figure 5:**
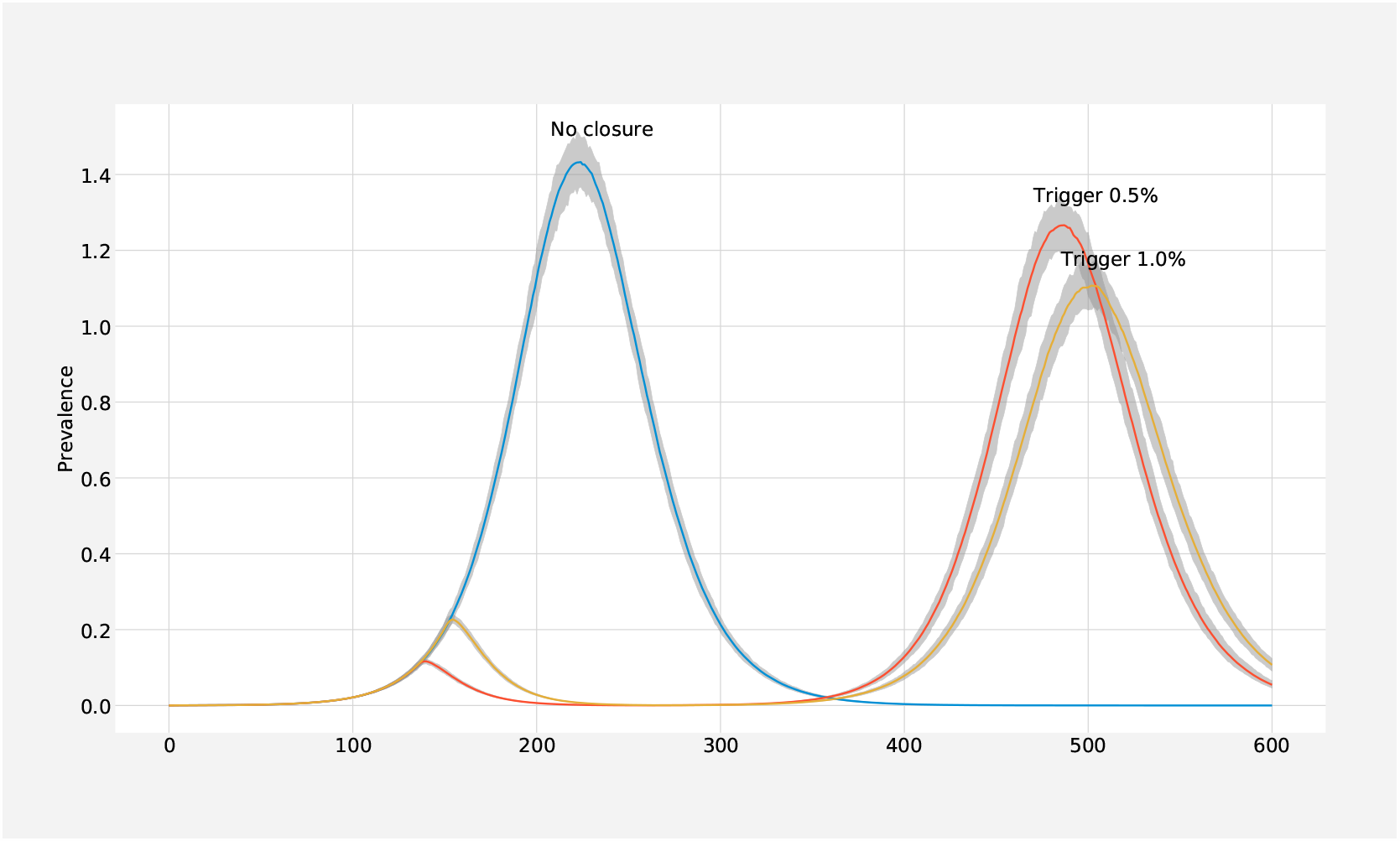
Comparison of triggers (1%and 0.5%) for social distancing policies with the no intervention strategy.

For an outbreak with baseline transmission rate our systematic and extensive simulations suggest the main benefit of social distancing is to buy time by delaying the peak as shown in Table 2. The length of social distancing phase has minimal to no effect on the magnitude of the second wave. However, it directly changes the timing of the second wave. This time until the second wave could help public health officials to prepare for a second wave. The threshold used to trigger the social distancing policy has significant implications on the epidemic dynamics. Specifically, early triggers result in a small first wave and a larger second wave. Conversely, later triggers result in a relatively bigger first wave, and smaller second wave when compared to the second waves of other policy triggering options. This result suggests that depending on the hospital bed and ICU capacity, early triggers or late triggers may be used to balance the magnitudes of the waves for mitigation and to manage the health care system capacity.

**Table 2:**
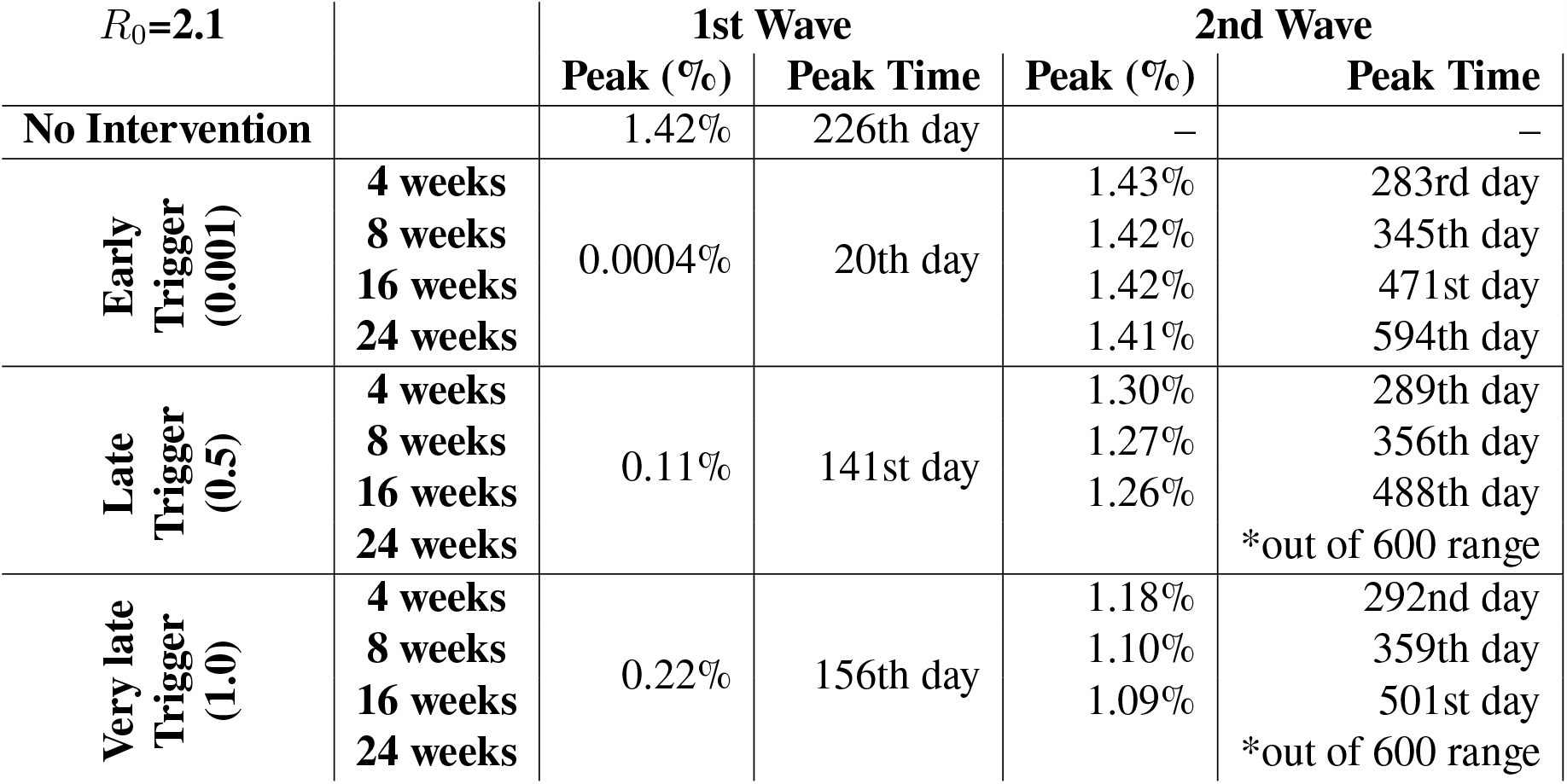
Total infection peak magnitude and peak timing with different school closure/ social distancing scenarios − no closure as baseline, 4 weeks closure, 8 weeks closure, 16 weeks closure, and 24 weeks closure.

The number of hospitalizations also follow a similar trend as preented in Figure 6 for social distancing. We compare hospitalization volumes for early and late trigger with 8 weeks and 16 weeks of social distancing. Using an earlier trigger results in a smaller initial peak in hospitalizations than triggering that later. Conversely, the second peak is observed higher when an early trigger is used than the later trigger. The length of the social distancing policy merely delays the second peak, both for early and late triggers, with no material impact to the size of the second peak. Using a smaller prevalence value for triggering the policy results in a smaller first peak than the later trigger, however, the second peak for early trigger is larger than that of late trigger. The length of the social distancing again affects the timing of the second peak.

**Figure 6:**
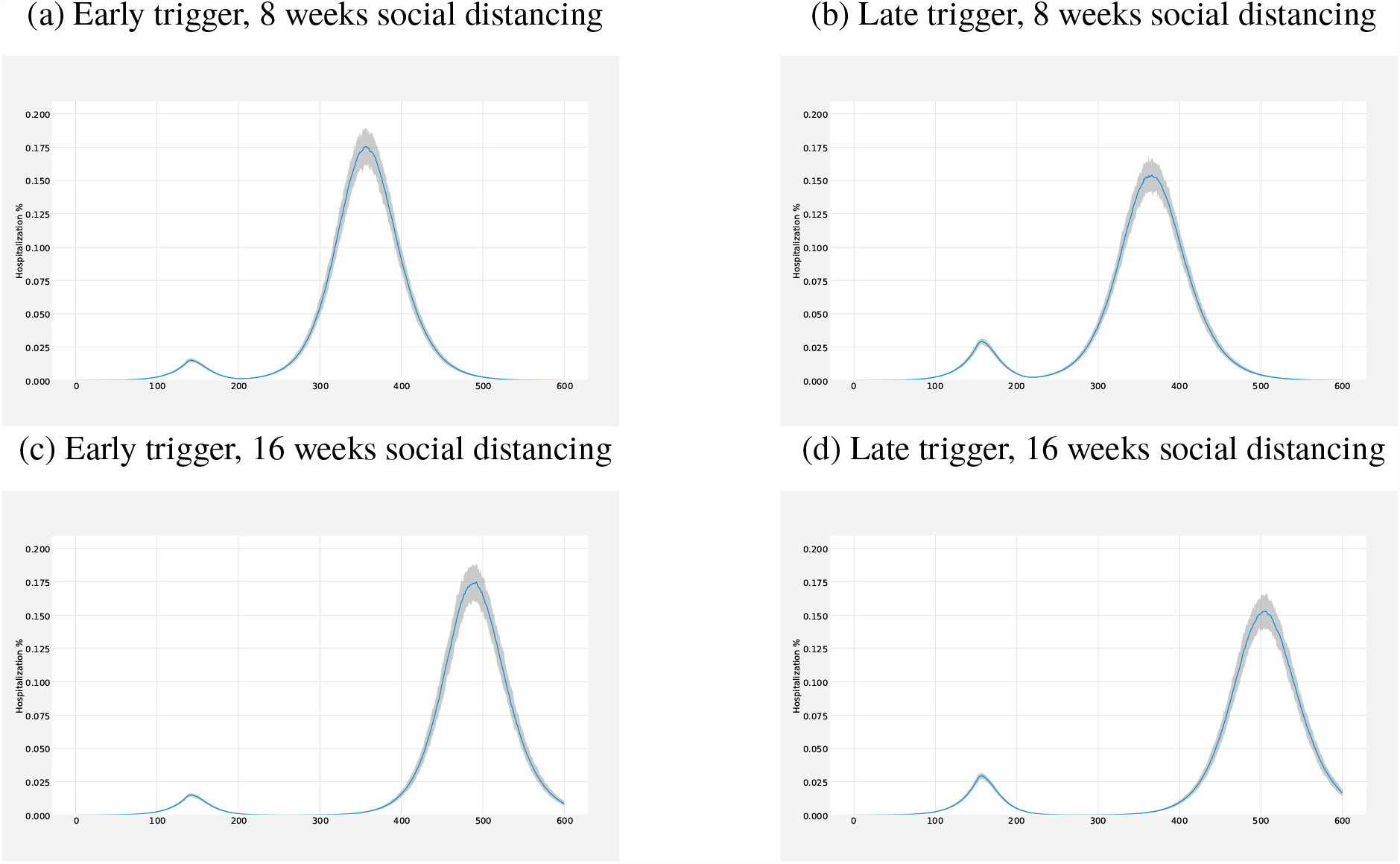
Comparison of hospitalization volumes for early and late trigger (i.e., columns) for 8 weeks and 16 weeks closures (i.e., rows).

### 4.2 Higher Transmission Scenario (*R*_0_ = 3.15)

While some highly dense US states observed faster growth in the number of cases, e.g., New York and California, some states with lower population density observed steep and later increase in the cases. Therefore, we simulate a scenario with higher transmission rate which is achieved with *R*_0_ = 3.15. Similar to base case scenario, simulations suggest the main benefit of social distancing is to buy time by delaying the peak as shown in Table 3. The length of social distancing phase has minimal to no effect on the magnitude, i.e., height of the peak of the second wave. However, it directly changes the timing of the second wave. The magnitudes of the second peaks for high transmission scenario, for all trigger options, is more than 2 times larger than those of for the baseline transmission scenario.

**Table 3:**
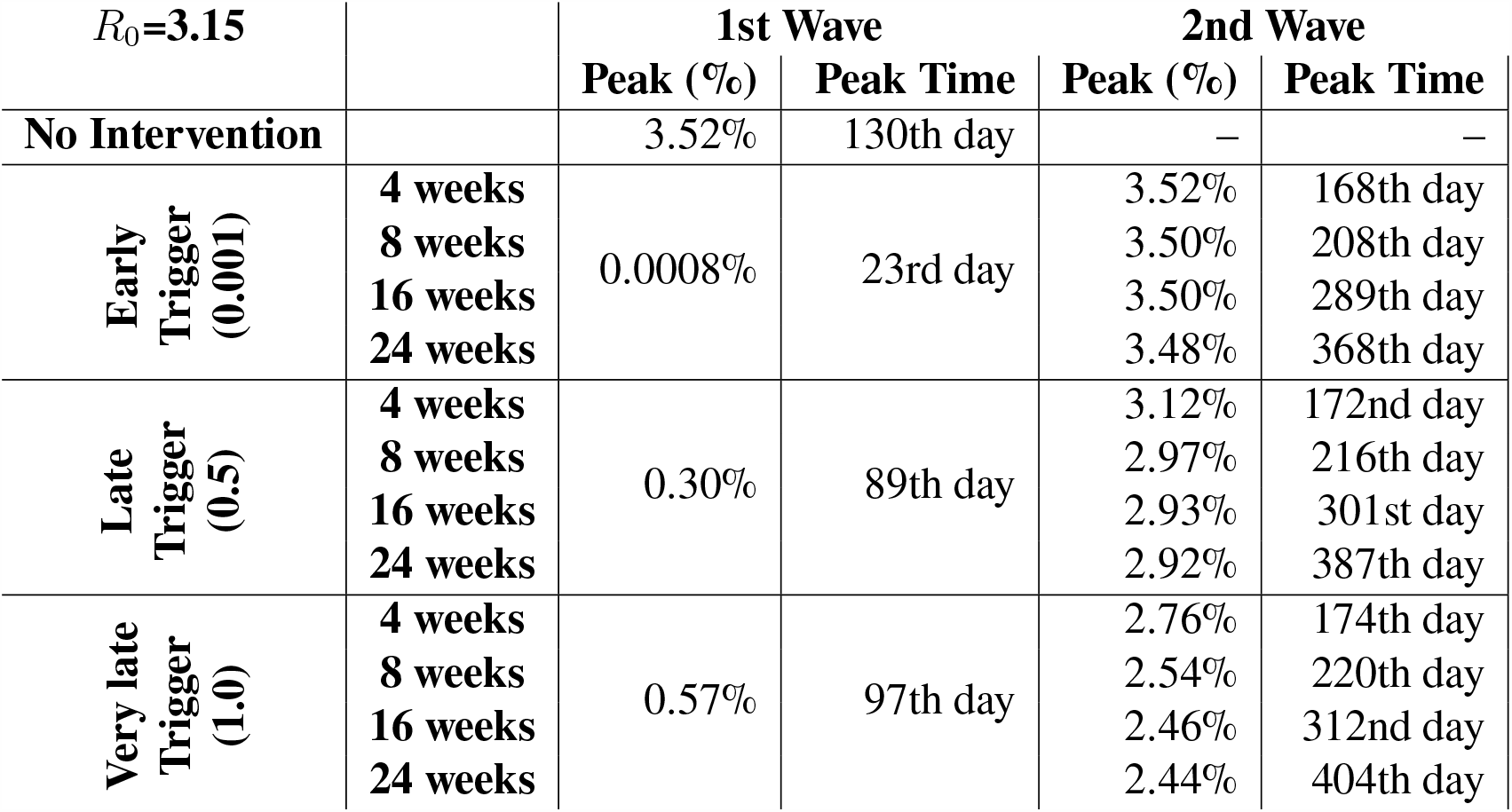
Total infection peak magnitude and peak timing with different school closure/ social distancing scenarios - no closure as baseline, 4 weeks closure, 8 weeks closure, 16 weeks closure, and 24 weeks closure.

Depending on the length of the social distancing implemented, public health officials gain time until the second wave ranging from one month, when the least aggressive scenario is employed, to 9 months, when the most aggressive scenario is employed. Such delay in the second peak could enable increasing hospital capacity, explorations for vaccinations and improved public health awareness.

### 4.3 Lower Transmission Scenario (*R*_0_ = 1.57)

For an outbreak with low transmission rate our simulations suggest the main benefit of social distancing, similar to baseline scenario and high transmission scenario, is to buy time by delaying the peak as shown in Table 4. However, for the low transmission scenario the magnitude of the peak, even with no intervention, is small compared to the high and baseline transmission rates. Furthermore, the timing of the peak for low transmission is much later than that of baseline transmission (i.e., 432nd day versus 226th day). In fact, the hospital capacity might be already enough or could be increased until the peak time to cover the 0.39% peak magnitude. Overall, the closure length should depend on the extra capacity needed to handle the patients during the peak.

**Table 4:** Total infection peak magnitude and peak timing with different social distancing scenarios - no closure as baseline, 4 weeks closure, 8 weeks closure, 16 weeks closure, and 24 weeks closure.

| $R_0=1.5$ | | 1st Wave | | 2nd Wave | |
| --- | --- | --- | --- | --- | --- |
|  |  | Peak (%) | Peak Time | Peak (%) | Peak Time |
| **No Intervention** |  | 0.39% | 432nd day | – | – |
| **Early Trigger**<br>(0.001) | **4 weeks** | 0.0003% | 19th day | 0.38% | 538th day |
|  | **8 weeks** |  |  |  | *out of 600 range |
|  | **16 weeks** |  |  |  | *out of 600 range |
|  | **24 weeks** |  |  |  | *out of 600 range |
| **Late Trigger**<br>(0.5) | **4 weeks** | 0.045% | 260th day | 0.34% | 551st day |
|  | **8 weeks** |  |  |  | *out of 600 range |
|  | **16 weeks** |  |  |  | *out of 600 range |
|  | **24 weeks** |  |  |  | *out of 600 range |
| **Very late Trigger**<br>(1.0) | **4 weeks** | 0.08% | 294th day | 0.30% | 556th day |
|  | **8 weeks** |  |  |  | *out of 600 range |
|  | **16 weeks** |  |  |  | *out of 600 range |
|  | **24 weeks** |  |  |  | *out of 600 range |

### 4.4 Evaluating Closure*/*Reopening Decisions

Since our compartmental model is age structured, it allows us to look at each age group separately in addition to overall population. This feature enables us to explore the vulnerability of high risk population. Therefore, here we compare cumulative attack rate (CAR), % deaths and peak hospitalizations in each age group separately for all transmission scenarios. In Figure 7, we present % deaths for each age group for different transmission scenario considered. We find that the burden of the pandemic on elderly population is the worse, in all potential transmission scenario. Specifically, as the transmission rate increases, cumulative death % for elderly population increases drastically. This suggests that for dense regions, high risk groups should be given more attention.

**Figure 7:**
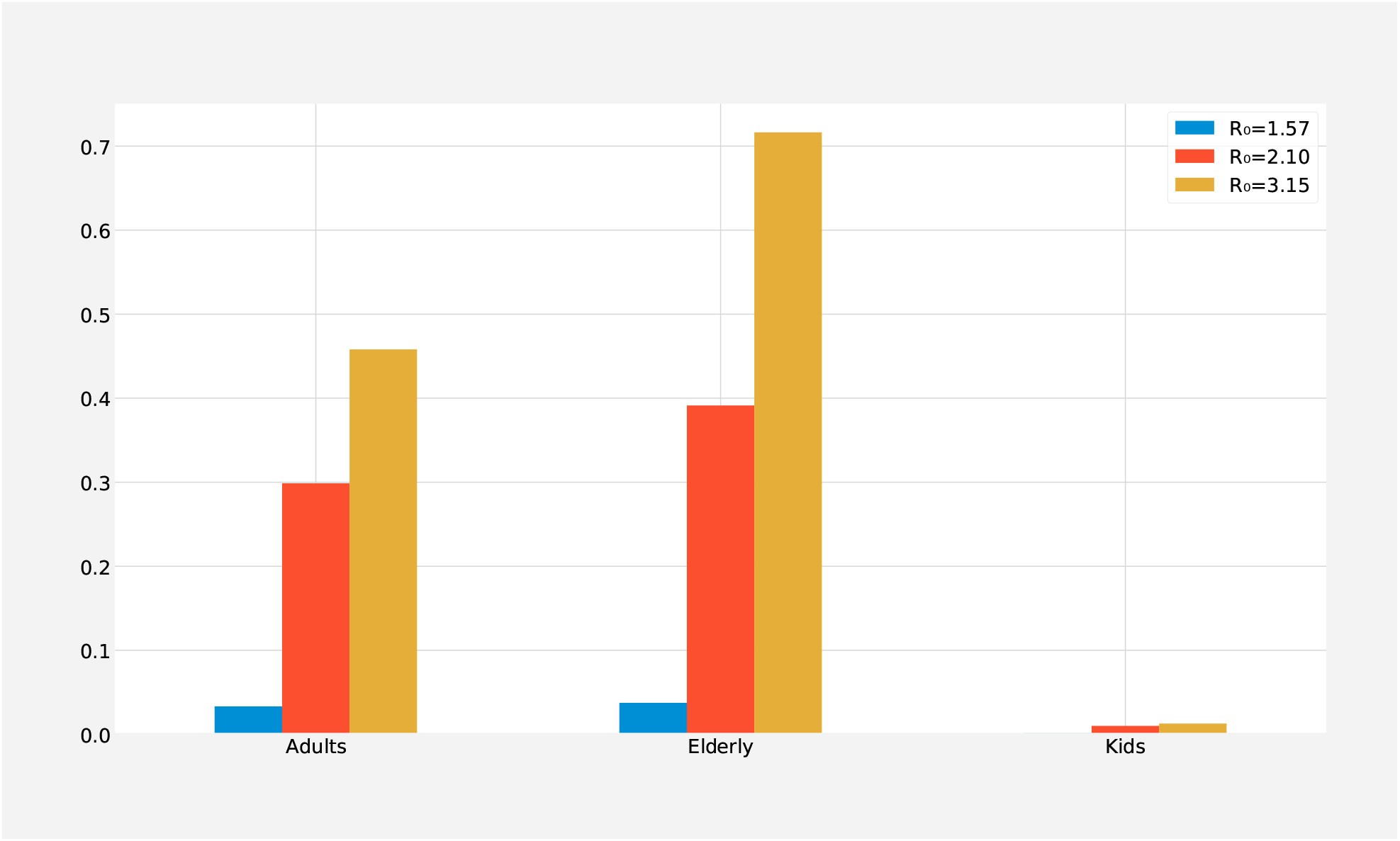
Cumulative Deaths (%) for different *R*_0_ values for each age group; social distancing is triggered when cumulative prevalence is reached 0.5% for 8 weeks.

On the other hand, early trigger and late trigger would result in different death tolls. We compare different closure strategies for all levels of social distancing along with different trigger levels (See Appendix C and Table 5). Early trigger results in a relatively lower death rate in low transmission rate while late trigger results in a larger death rate as shown in Table 5. This result is presented for 16 weeks of social distancing. However, for the base scenario and higher transmission rate scenario the expected deaths would be lower when later trigger levels are used. Appendix C shows that till the 16 week of social distancing (1 week, 2 weeks, 3 weeks, 4 weeks and 8 weeks) in all transmission rates death % decreases as we increase the trigger level.(See Appendix C and Tables 10, 11, 12, 13, 14, 15).

**Table 5:** Death rate, maximum hospitalization rate, Cumulative attack rate (CAR) and Cumulative attack rate including the asymptomatic cases with 16 weeks of social distancing.

| $R_0$ | Trigger % | Death(%) | Peak Hospitalization Rate | CAR | CAR (+asymptomatic) |
| --- | --- | --- | --- | --- | --- |
| 1.57 | - | 0.135 | 0.051 | 18.203 | 22.191 |
| 1.57 | 0.001 | 0.0003 | 0.0001 | 0.048 | 0.060 |
| 1.57 | 0.5 | 0.006 | 0.005 | 0.837 | 1.020 |
| 1.57 | 1.0 | 0.012 | 0.010 | 1.613 | 1.966 |
| 2.1 | - | 0.270 | 0.197 | 33.587 | 40.914 |
| 2.1 | 0.001 | 0.268 | 0.197 | 33.453 | 40.812 |
| 2.1 | 0.5 | 0.261 | 0.174 | 32.618 | 39.797 |
| 2.1 | 1.0 | 0.251 | 0.153 | 31.550 | 38.450 |
| 3.15 | - | 0.438 | 0.522 | 49.125 | 59.829 |
| 3.15 | 0.001 | 0.437 | 0.521 | 49.147 | 59.865 |
| 3.15 | 0.5 | 0.428 | 0.436 | 48.147 | 58.650 |
| 3.15 | 1.0 | 0.421 | 0.365 | 47.240 | 57.547 |

To control the rapidly growing outbreak across the US, public health officials and governments issued shelter-in-place orders. Although social distancing and shelter-in-place orders help mitigate the outbreak, it has significant economic and social costs. For example, in the US during the social distancing for COVID-19, the unemployment rates have spiked. To identify possible reopening strategies, we evaluate a two-phase reopening strategy. We compare this strategy to two basic alternatives: (1) No closures (i.e., no social distancing), (2) Immediate reopening (i.e., no phases).

For example, Figure 8 shows the three different scenarios: (1) *no-closure*, (2) *Immediate reopening*, and (3) *Reopening with Phases* for an outbreak with medium growth rate. The no-closure strategy is without any social distancing implemented, indicated by the red line. The immediate reopening strategy is returning back to original contact rates for all age groups immediately after 8 weeks of closure shown by the yellow line. The reopening with phases strategy, indicated by the blue line in Figure 8a, corresponds to reopening with phases of length 8-weeks, i.e., after 8 weeks of social distancing an 8-week long phase 1 starts with increases the contact rate with certain rate. Phase 1 is followed by another 8-week long phase 2, where contact rates are even more increased. Finally, after phase 2, original contact rates are restored (See Appendix Table 8 for the corresponding contact rates). In Figure 8a, we observe that phased reopening results in smaller outbreak sizes (i.e., smaller peak magnitude) than both the no-closure strategy and the immediate reopening strategy. We have also evaluated reopening strategies with 24-week length phases. Figure 8b shows that the burden of the pandemic is further mitigated using longer term phases.

**Figure 8:**
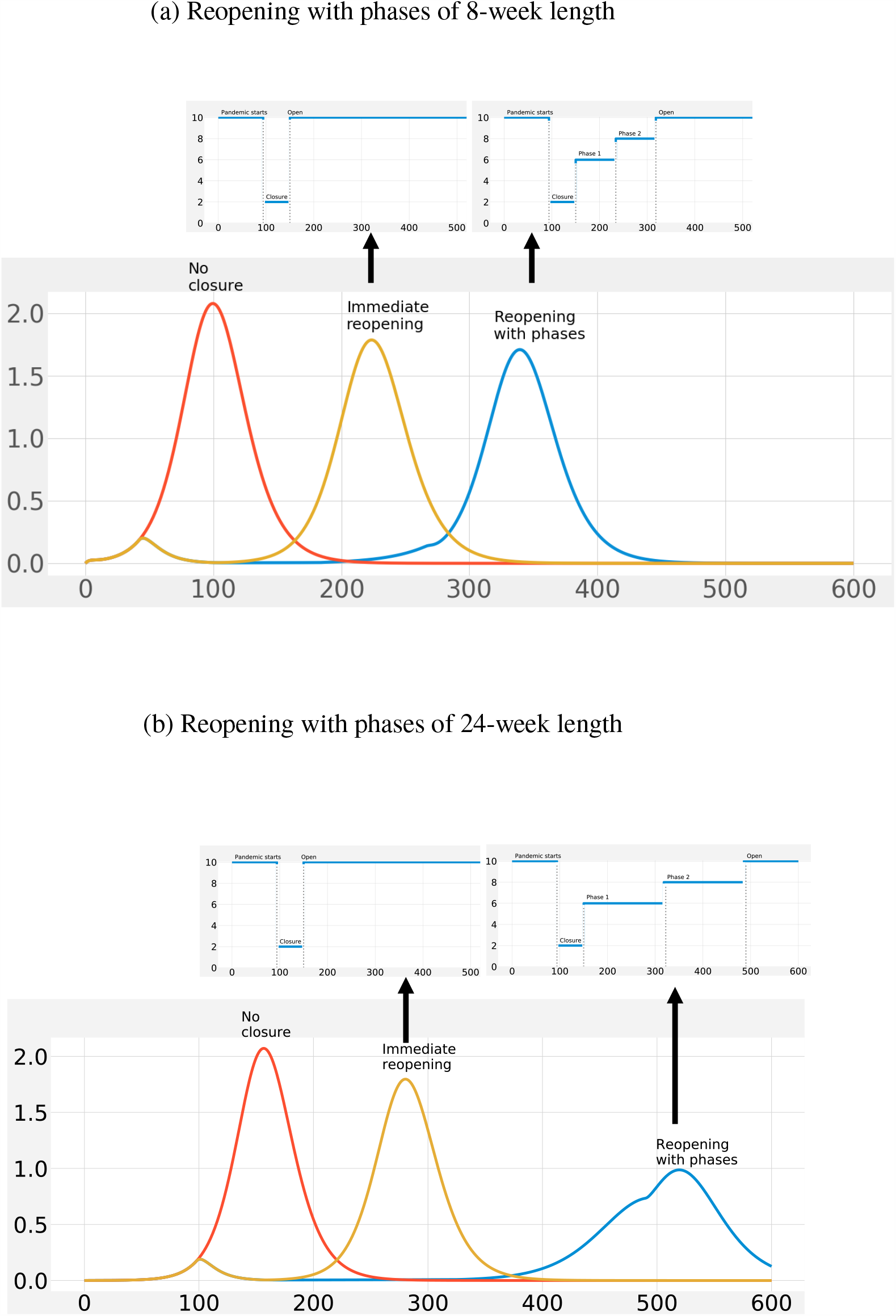
Prevalence curve comparison in ”No closure”, ”Immediate reopening” and ”Reopening with Phases” for medium growth rate. (a) 8 weeks reopening strategy, (b) 24 weeks reopening strategy

Figure 8 shows the different scenarios for the base transmission scenario and two possible lengths of reopening phases. We also show outbreaks with different growth rate and with other possible lengths of reopening phases. For example, Figure 9 shows different possible lengths of reopening phases for an outbreak with baseline transmission rate. For all lengths considered, the longer the phase duration is, the more dampened the outbreak burden, as well as the further in time the peak is observed. For example, the peak is reduced more than half with 24-week long phases instead of 4-week long phases. This presents a trade-off between hospital capacity especially how much it could be enhanced with the delay of the peak by social distancing and the size of the second peak as a result of the length of social distancing.

**Figure 9:**
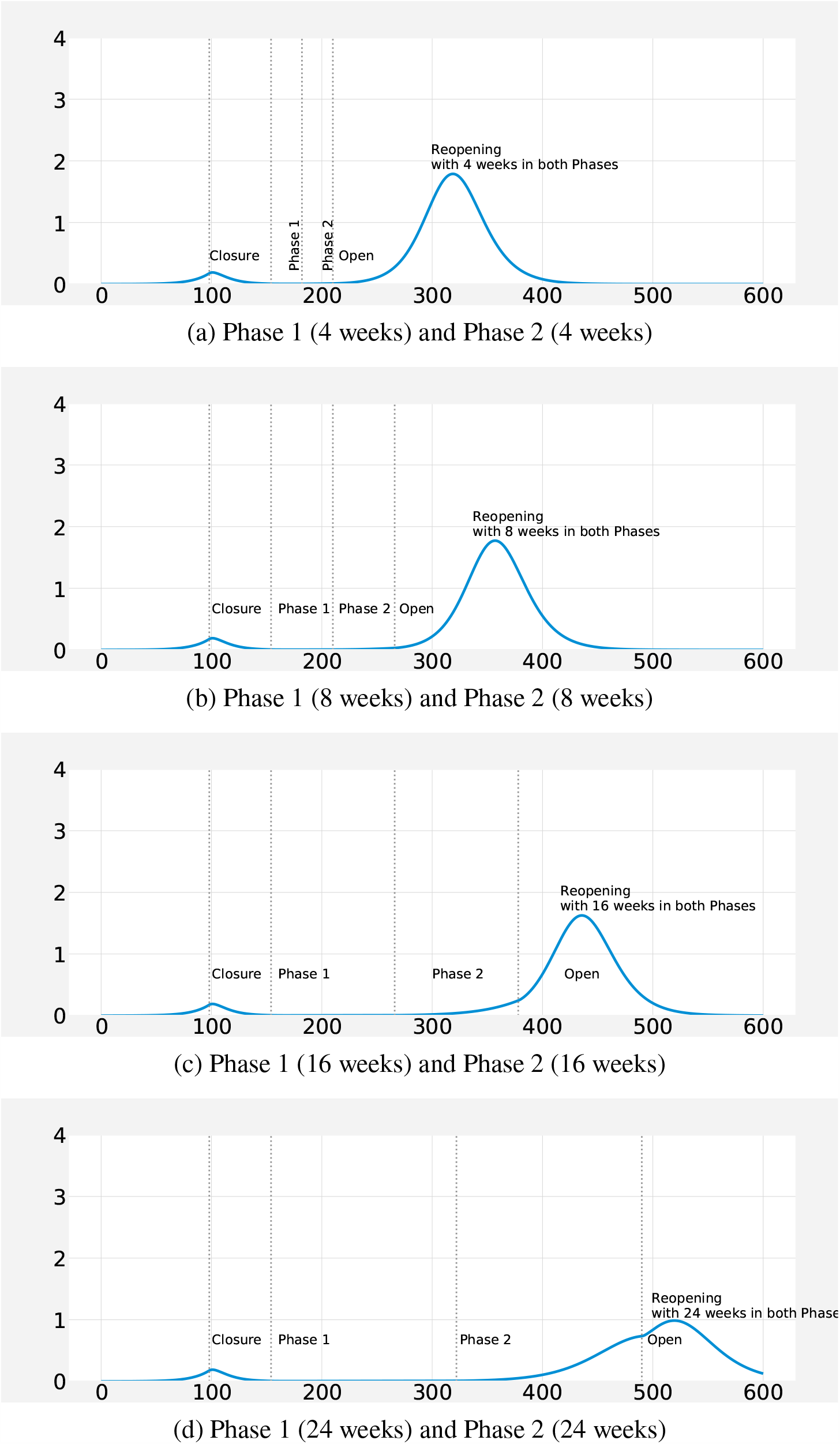
Comparison of the different reopening strategies for *R*_0_: 2.1, 8 weeks of closure followed by (a) 4 weeks in Phase 1 and Phase 2, (b) 8 weeks in Phase 1 and Phase 2, (c) 16 weeks in Phase 1 and Phase 2 (d) 24 weeks in Phase 1 and Phase 2.

Similarly, Figure 10 shows different possible lengths of reopening phases for an outbreak with higher transmission case. Again, for all lengths of social distancing considered, the longer the phase windows the more the reduction in the epidemic size is achieved. Though, surprisingly, the observed peak time for different reopening phases shifts marginally.

**Figure 10:**
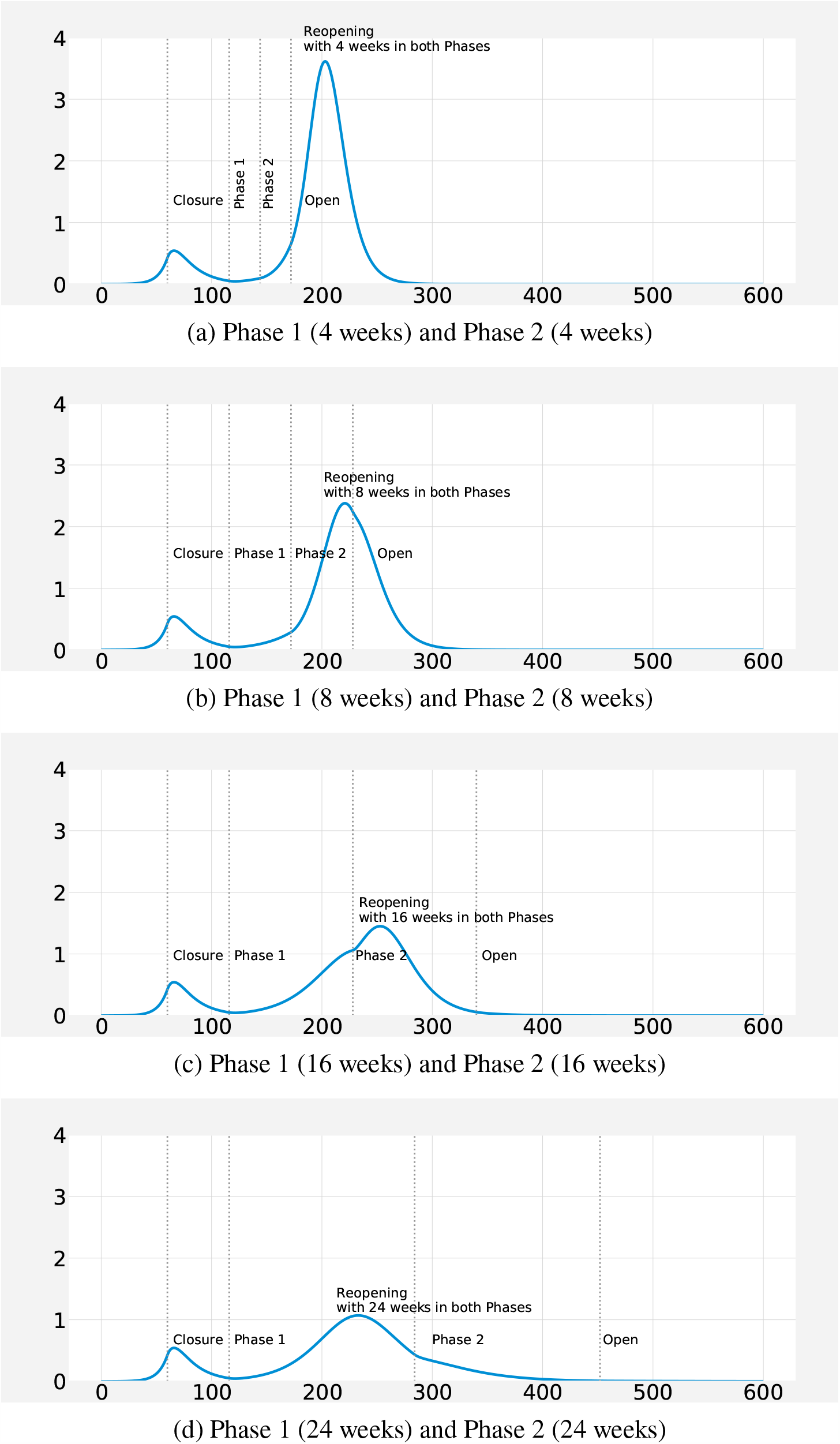
Comparison of the different reopening strategies for *R*_0_: 3.15, 8 weeks of closure followed by (a) 4 weeks in Phase 1 and Phase 2, (b) 8 weeks in Phase 1 and Phase 2, (c) 16 weeks in Phase 1 and Phase 2 (d) 24 weeks in Phase 1 and Phase 2.

Finally, Figure 11 shows different possible duration of reopening phases for the pandemic with smaller growth rate. Unsurprisingly, the overall outbreak is much smaller. Specific to this scenario, reopening phases with 16-week and 24-week length of closure delays the peak timing even beyond the next 600 days. Arguably, the healthcare system could absorb a reopening with the minimum length considered, i.e., 4-weeks. In other words, we note that for regions with low transmission rate with enough hospital capacity, reopening relatively sooner might be a feasible strategy.

**Figure 11:**
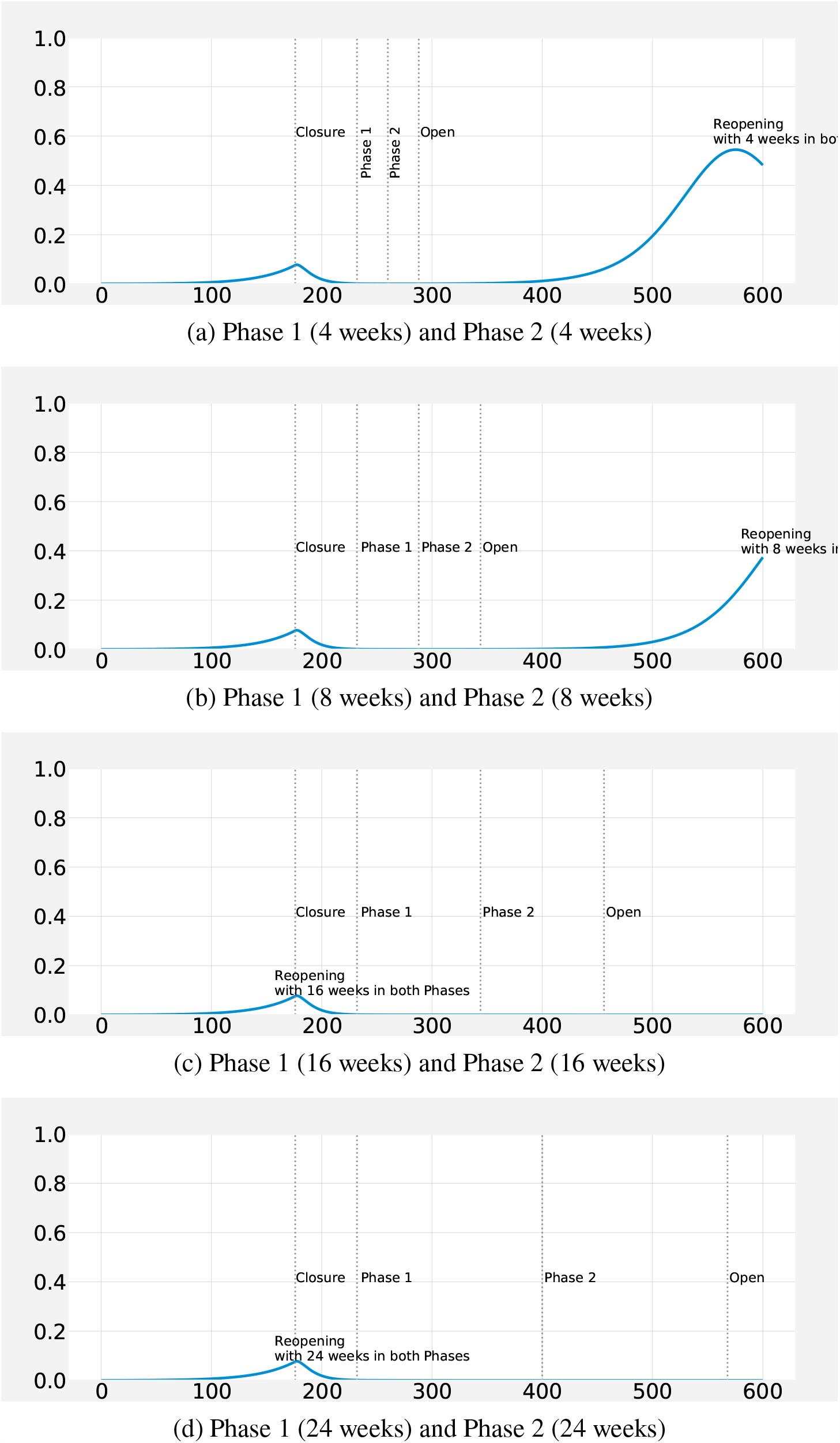
Comparison of the different reopening strategies for *R*_0_: 1.57, 8 weeks of closure followed by (a) 4 weeks in Phase 1 and Phase 2, (b) 8 weeks in Phase 1 and Phase 2, (c) 16 weeks in Phase 1 and Phase 2 (d) 24 weeks in Phase 1 and Phase 2.

Table 6 summarizes possible scenarios that minimizes expected Cumulative Attack Rate (CAR) and expected total death rate under different transmission cases considered. For lower transmission rate, an 8-week closure results in a significantly bigger reduction in CAR as well as total death toll than 4-week closure. For medium transmission rate, i.e., the baseline, we observe that a 24-week closure results in a significantly higher reduction in CAR as well as total death toll. For higher transmission rate, although a 24-week closure results in highest reduction in CAR as well as total death toll, the overall reduction magnitudes are very small. Furthermore, the marginal reduction due to additional weeks to closure appear to be negligible. To achieve reductions similar to other transmission rates longer closure scenarios should be considered.

**Table 6:** Highlight of different scenarios (i.e., different closure triggers and lengths) for the three transmission rates that minimizes both the cumulative attack rate (CAR) and total death toll, separately. The bold rows indicate the scenarios for each transmission rate, that minimizes these objectives. Note that independently minimizing CAR and minimizing total deaths result in the same scenarios.

|  | Objectives |  |  |  |  |  |
| --- | --- | --- | --- | --- | --- | --- |
|  | Min. CAR |  |  | Min. Death |  |  |
|  | Threshold | Closure | Reduction (%) | Threshold | Closure | Reduction (%) |
| $R_0 = 1.57$ | 1% | 4 weeks | -0.296 | 1% | 4 weeks | -0.326 |
| $R_0 = \mathbf{1.57}$ | **1%** | **8 weeks** | **-0.779** | **1%** | **8 weeks** | **-0.791** |
| $R_0 = 1.57$ | | | *out of 600 day range | | | |
| $R_0 = 1.57$ | | | *out of 600 day range | | | |
| $R_0 = 2.10$ | 1% | 4 weeks | -0.029 | 1% | 4 weeks | -0.030 |
| $R_0 = 2.10$ | 1% | 8 weeks | -0.040 | 1% | 8 weeks | -0.041 |
| $R_0 = 2.10$ | 1% | 16 weeks | -0.061 | 1% | 16 weeks | -0.070 |
| $R_0 = \mathbf{2.10}$ | **1%** | **24 weeks** | **-0.749** | **1%** | **24 weeks** | **-0.781** |
| $R_0 = 3.15$ | 1% | 4 weeks | -0.027 | 1% | 4 weeks | -0.027 |
| $R_0 = 3.15$ | 1% | 8 weeks | -0.033 | 1% | 8 weeks | -0.034 |
| $R_0 = 3.15$ | 1% | 16 weeks | -0.038 | 1% | 16 weeks | -0.037 |
| $R_0 = \mathbf{3.15}$ | **1%** | **24 weeks** | **-0.041** | **1%** | **24 weeks** | **-0.041** |

**Table 7:**
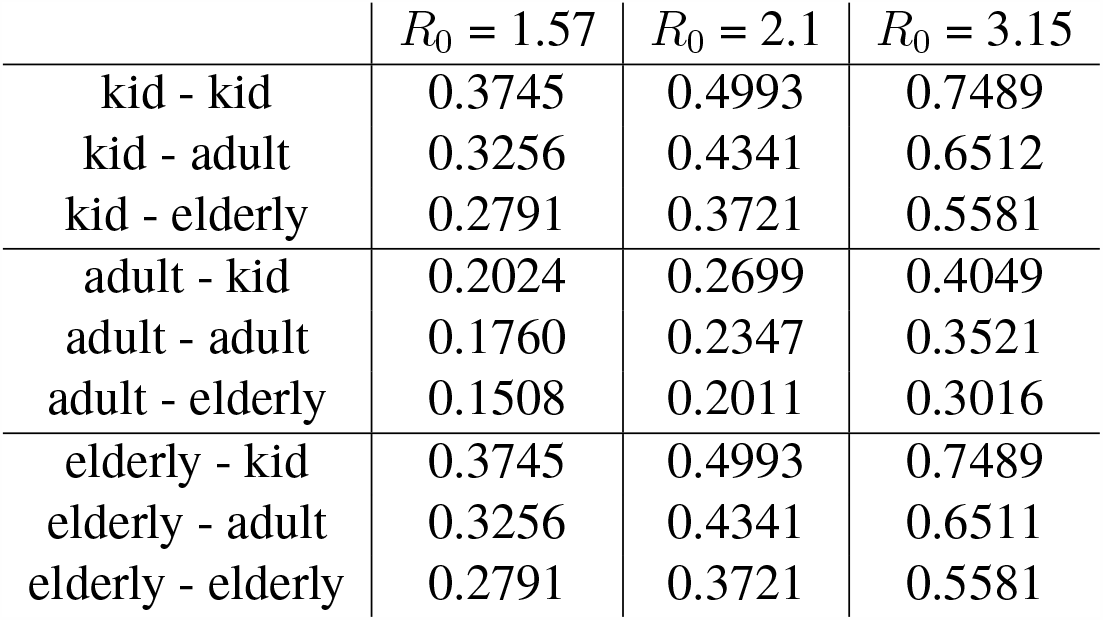
Age specific transmission rate (*β*) for different reproduction numbers (*R*_0_).

| | $R_0 = 1.57$ | $R_0 = 2.1$ | $R_0 = 3.15$ |
| --- | --- | --- | --- |
| kid - kid | 0.3745 | 0.4993 | 0.7489 |
| kid - adult | 0.3256 | 0.4341 | 0.6512 |
| kid - elderly | 0.2791 | 0.3721 | 0.5581 |
| adult - kid | 0.2024 | 0.2699 | 0.4049 |
| adult - adult | 0.1760 | 0.2347 | 0.3521 |
| adult - elderly | 0.1508 | 0.2011 | 0.3016 |
| elderly - kid | 0.3745 | 0.4993 | 0.7489 |
| elderly - adult | 0.3256 | 0.4341 | 0.6511 |
| elderly - elderly | 0.2791 | 0.3721 | 0.5581 |

**Table 8:** Age specific contact rates (*c*_*ij*_) for open condition - no social distancing, closed condition - social distancing, Reopening condition - Phase 1 and Reopening condition - Phase 2.

|  | Open Cond. | Closed Cond. | Phase 1 - Reopening Cond. | Phase 2 - Reopening Cond. |
| --- | --- | --- | --- | --- |
| kid - kid | 0.63 | 0.13 | 0.38 | 0.50 |
| kid - adult | 0.31 | 0.19 | 0.25 | 0.28 |
| kid - elderly | 0.06 | 0.06 | 0.06 | 0.06 |
| adult - kid | 0.38 | 0.13 | 0.25 | 0.31 |
| adult - adult | 0.50 | 0.19 | 0.38 | 0.44 |
| adult - elderly | 0.13 | 0.06 | 0.06 | 0.13 |
| elderly - kid | 0.06 | 0.13 | 0.06 | 0.06 |
| elderly - adult | 0.63 | 0.31 | 0.47 | 0.53 |
| elderly - elderly | 0.31 | 0.31 | 0.31 | 0.31 |

**Table 9:** Age specific hospitalization rate, recovery rate and mortality rate.

|  | Hospitalization Rate | Recovery Rate | Mortality Rate |
| --- | --- | --- | --- |
| Kid | 0.01 - 0.05 | 0.999 | 0.001 |
| Adult | 0.22 - 0.26 | 0.986 | 0.014 |
| Elderly | 0.60 - 0.80 | 0.935 | 0.065 |

**Table 10:**
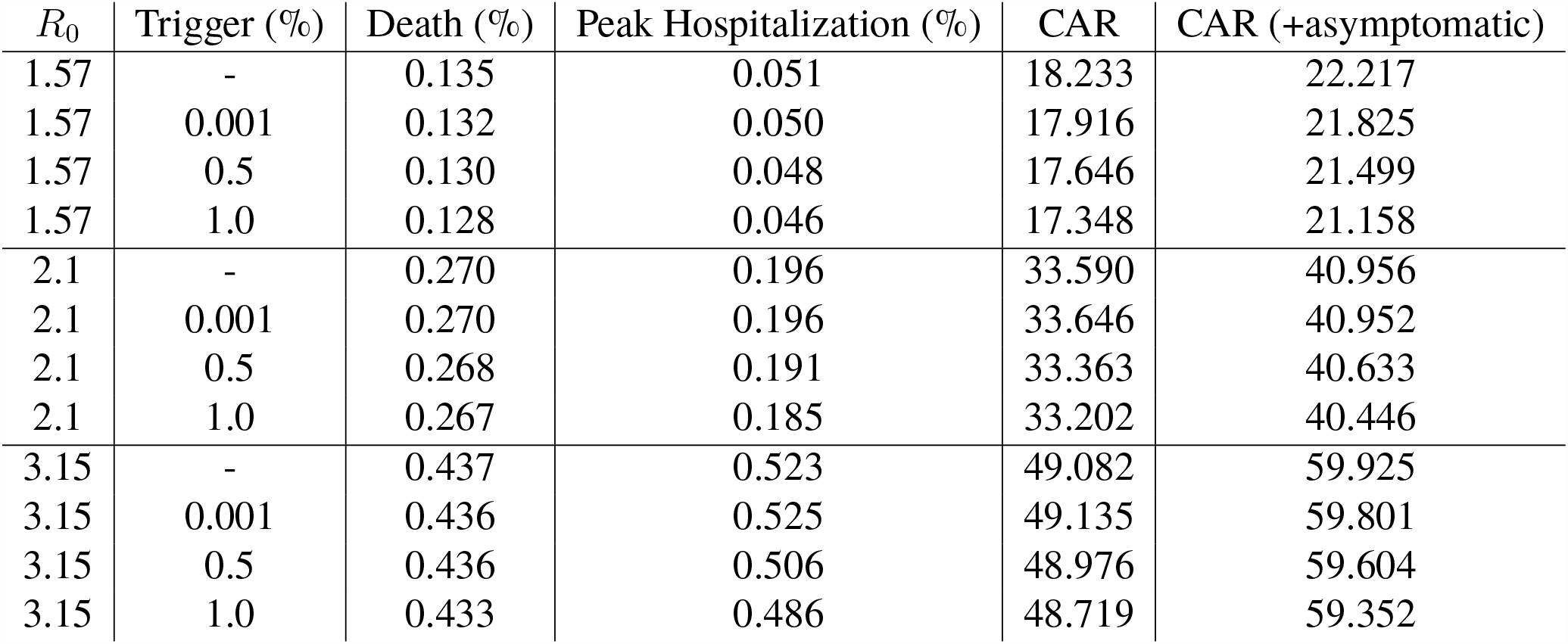
Death rate, maximum hospitalization rate, Cumulative attack rate (CAR) and Cumulative attack rate including the asymptomatic cases with 1 week of social distancing.

**Table 11:**
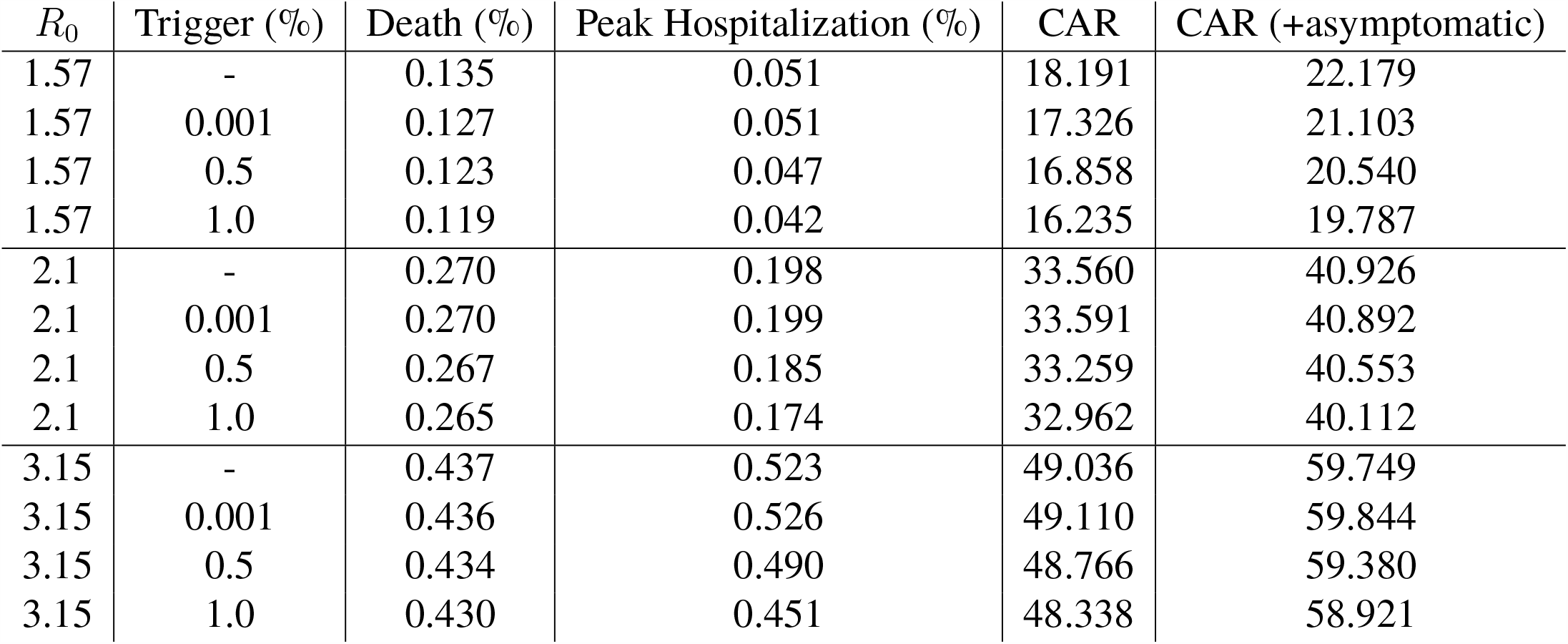
Death rate, maximum hospitalization rate, Cumulative attack rate (CAR) and Cumulative attack rate including the asymptomatic cases with 2 weeks of social distancing.

| $R_0$ | Trigger (%) | Death (%) | Peak Hospitalization (%) | CAR | CAR (+asymptomatic) |
| --- | --- | --- | --- | --- | --- |
| 1.57 | - | 0.135 | 0.051 | 18.191 | 22.179 |
| 1.57 | 0.001 | 0.127 | 0.051 | 17.326 | 21.103 |
| 1.57 | 0.5 | 0.123 | 0.047 | 16.858 | 20.540 |
| 1.57 | 1.0 | 0.119 | 0.042 | 16.235 | 19.787 |
| 2.1 | - | 0.270 | 0.198 | 33.560 | 40.926 |
| 2.1 | 0.001 | 0.270 | 0.199 | 33.591 | 40.892 |
| 2.1 | 0.5 | 0.267 | 0.185 | 33.259 | 40.553 |
| 2.1 | 1.0 | 0.265 | 0.174 | 32.962 | 40.112 |
| 3.15 | - | 0.437 | 0.523 | 49.036 | 59.749 |
| 3.15 | 0.001 | 0.436 | 0.526 | 49.110 | 59.844 |
| 3.15 | 0.5 | 0.434 | 0.490 | 48.766 | 59.380 |
| 3.15 | 1.0 | 0.430 | 0.451 | 48.338 | 58.921 |

**Table 12:** Death rate, maximum hospitalization rate, Cumulative attack rate (CAR) and Cumulative attack rate including the asymptomatic cases with 3 weeks of social distancing.

| $R_0$ | Trigger (%) | Death (%) | Peak Hospitalization (%) | CAR | CAR (+asymptomatic) |
| --- | --- | --- | --- | --- | --- |
| 1.57 | - | 0.135 | 0.051 | 18.219 | 22.226 |
| 1.57 | 0.001 | 0.119 | 0.050 | 16.375 | 19.975 |
| 1.57 | 0.5 | 0.113 | 0.045 | 15.563 | 18.964 |
| 1.57 | 1.0 | 0.106 | 0.040 | 14.710 | 17.945 |
| 2.1 | - | 0.270 | 0.198 | 33.580 | 40.886 |
| 2.1 | 0.001 | 0.270 | 0.197 | 33.613 | 40.932 |
| 2.1 | 0.5 | 0.266 | 0.181 | 33.113 | 40.350 |
| 2.1 | 1.0 | 0.262 | 0.167 | 32.637 | 39.777 |
| 3.15 | - | 0.437 | 0.518 | 49.106 | 59.807 |
| 3.15 | 0.001 | 0.437 | 0.522 | 49.194 | 59.895 |
| 3.15 | 0.5 | 0.432 | 0.472 | 48.593 | 59.224 |
| 3.15 | 1.0 | 0.428 | 0.431 | 48.096 | 58.555 |

**Table 13:** Death rate, maximum hospitalization rate, Cumulative attack rate (CAR) and Cumulative attack rate including the asymptomatic cases with 4 weeks of social distancing.

| $R_0$ | Trigger (%) | Death (%) | Peak Hospitalization (%) | CAR | CAR (+asymptomatic) |
| --- | --- | --- | --- | --- | --- |
| 1.57 | - | 0.135 | 0.050 | 18.222 | 22.188 |
| 1.57 | 0.001 | 0.108 | 0.050 | 15.008 | 18.266 |
| 1.57 | 0.5 | 0.098 | 0.045 | 13.696 | 16.724 |
| 1.57 | 1.0 | 0.091 | 0.039 | 12.821 | 15.606 |
| 2.1 | - | 0.270 | 0.196 | 33.592 | 40.957 |
| 2.1 | 0.001 | 0.270 | 0.196 | 33.575 | 40.935 |
| 2.1 | 0.5 | 0.266 | 0.179 | 33.098 | 40.269 |
| 2.1 | 1.0 | 0.262 | 0.163 | 32.611 | 39.688 |
| 3.15 | - | 0.437 | 0.524 | 49.156 | 59.853 |
| 3.15 | 0.001 | 0.436 | 0.523 | 49.059 | 59.800 |
| 3.15 | 0.5 | 0.431 | 0.463 | 48.428 | 59.070 |
| 3.15 | 1.0 | 0.425 | 0.413 | 47.832 | 58.281 |

**Table 14:** Death rate, maximum hospitalization rate, Cumulative attack rate (CAR) and Cumulative attack rate including the asymptomatic cases with 8 weeks of social distancing.

| $R_0$ | Trigger (%) | Death (%) | Peak Hospitalization (%) | CAR | CAR (+asymptomatic) |
| --- | --- | --- | --- | --- | --- |
| 1.57 | - | 0.134 | 0.051 | 18.163 | 22.167 |
| 1.57 | 0.001 | 0.032 | 0.036 | 4.866 | 5.948 |
| 1.57 | 0.5 | 0.028 | 0.026 | 4.175 | 5.096 |
| 1.57 | 1.0 | 0.028 | 0.018 | 4.010 | 4.903 |
| 2.1 | - | 0.270 | 0.197 | 33.579 | 40.923 |
| 2.1 | 0.001 | 0.270 | 0.198 | 33.567 | 40.933 |
| 2.1 | 0.5 | 0.265 | 0.176 | 32.984 | 40.182 |
| 2.1 | 1.0 | 0.259 | 0.154 | 32.243 | 39.287 |
| 3.15 | - | 0.437 | 0.524 | 49.054 | 59.887 |
| 3.15 | 0.001 | 0.436 | 0.521 | 49.053 | 59.827 |
| 3.15 | 0.5 | 0.429 | 0.444 | 48.241 | 58.764 |
| 3.15 | 1.0 | 0.422 | 0.377 | 47.449 | 57.760 |

**Table 15:** Death rate, maximum hospitalization rate, Cumulative attack rate (CAR) and Cumulative attack rate including the asymptomatic cases with 24 weeks of social distancing.

| $R_0$ | Trigger (%) | Death (%) | Peak Hospitalization (%) | CAR | CAR (+asymptomatic) |
| --- | --- | --- | --- | --- | --- |
| 1.57 | - | 0.135 | 0.051 | 18.219 | 22.182 |
| 1.57 | 0.001 | 0.001 | 0.001 | 0.003 | 0.005 |
| 1.57 | 0.5 | 0.006 | 0.005 | 0.810 | 0.987 |
| 1.57 | 1.0 | 0.011 | 0.010 | 1.590 | 1.938 |
| 2.1 | - | 0.270 | 0.197 | 33.553 | 40.928 |
| 2.1 | 0.001 | 0.134 | 0.196 | 19.309 | 23.614 |
| 2.1 | 0.5 | 0.083 | 0.147 | 12.357 | 15.083 |
| 2.1 | 1.0 | 0.059 | 0.083 | 8.412 | 10.262 |
| 3.15 | - | 0.437 | 0.523 | 49.098 | 59.806 |
| 3.15 | 0.001 | 0.437 | 0.519 | 49.147 | 59.825 |
| 3.15 | 0.5 | 0.427 | 0.434 | 48.016 | 58.597 |
| 3.15 | 1.0 | 0.419 | 0.360 | 47.149 | 57.479 |

As for reopening, we also show that reopening with phases might be more crucial in dense populations like New York where higher transmission is estimated. Especially, long time window reopening can significantly reduce the burden of the outbreak; which makes it easier for the healthcare system. However, in places where lower transmission rates are estimated, e.g., in Nebraska, reopening with phases will also be useful but as critical as in New York. Even the shortest time-window reopening strategies considered results with cases that the healthcare system can absorb. To summarize, *one size fits all* reopening policies treating entire population as one is vulnerable to result in inefficiencies. Premature reopening strategies may result in excessive overload in hospitalizations and ICU capacity creating a deadlock in healthcare system. Reopening strategies with phases might provide a feasible middle ground to manage the trade-off between improving the economy and giving more time to health officials to improve healthcare capacity and develop vaccines.

Early reopening might result in increased number of infections and death tolls in the dense areas and late reopening might result in economic loss in comparatively less dense areas. Each region should consider its own hospital/ICU capacity and other limitations while easing social distancing. Our model provides a framework to consider the various trade-offs (e.g., the length of social distancing vs the number of hospitalizations) at a local level.

## 5 Conclusions

In this paper, we present a compartmental model for COVID-19 outbreak dynamics with three age groups, (i.e. kids, adults and elderly). We perform systematic and extensive simulations to evaluate social distancing and reopening scenarios for regions with different disease dynamics. Specifically, we find that the effective growth rate of the disease is different for different regions in the US. For example, New York, i.e., high density urban population, has a higher transmission rate whereas Nebraska, i.e., low density more rural population, has a lower transmission rate than the base case considered. Our simulations suggest that depending on the transmission rate estimated, the magnitude and the timing of the peak cases would vary across regions; thus the optimal mitigation policies may vary.

Furthermore, we present a trade-off between overall death toll of the outbreak and the enhanced hospital capacity in a region by preparations during the delayed time of the outbreak peak. Specifically, we consider triggering social distancing early (0.001%), late (0.5%), or very late (1%). We find that early trigger social distancing strategies result in small death tolls, however relatively larger second waves. Conversely, late trigger social distancing strategies result in higher initial death tolls but relatively small second waves This study show that policy makers should expect multiple waves of cases as a result of the social distancing policies implemented when there are no vaccines available for mass immunization and appropriate antiviral treatments. Social distancing policies only provide time for managing the cases in the population until these pharmaceutical interventions become available. We have shown that with higher transmission scenario it is better to use relatively later trigger with longer closure duration. Also our results show that social distancing is comparatively more effective when there is lower transmission in terms of the time gained for pharmaceutical interventions mentioned above.

Finally, we note that there is so much unknown about COVID-19, and our models include several assumptions regarding parameter values as summarized in Table 1. Although, we expect the relative comparisons of different strategies should still apply, the results of this work should be considered within the limitations of the model parameters. Our results are based on parameters obtained mainly from CDC reports and early estimates of the disease. As the parameters are refined, our projections will be improved. The number of hospitalizations and deaths will also depend on how effectively we protect our high risk populations. In the reopening scenarios, phase 1 and phase 2 are solely based on contact rate relaxations in social distancing. Depending on the current situation, the restrictions and contact rates may have been tightened to prevent hospitalizations for overwhelming local capacities. As a future work, we are planning on controlling the spread proactively so that we could control the hospital capacity and actively use the risk-based guidelines to manage the pandemic speed.

## Data Availability

Data is available to the public

## A Epidemic Model Parameters

## B Basic Reproduction Number 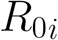

The basic reproductive number 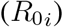 is define as the number of secondary cases an infectious individual on a particular age group *i* generates during the time period he/she is infectious on the susceptible population of their age group at the beginning of an epidemic. The individuals that can potentially infect the population in our model are infected individuals either symptomatic or asymptomatic in this case. To compute the 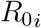 we use the next generation operator Van den Driessche & Watmough (2002). Let the vector *F* be the rate of new infections flowing to the latent compartment and the vector *V* to be the rate of transfer of individuals out of the compartment that are able to transmit the disease. Then using our system of equations we define:

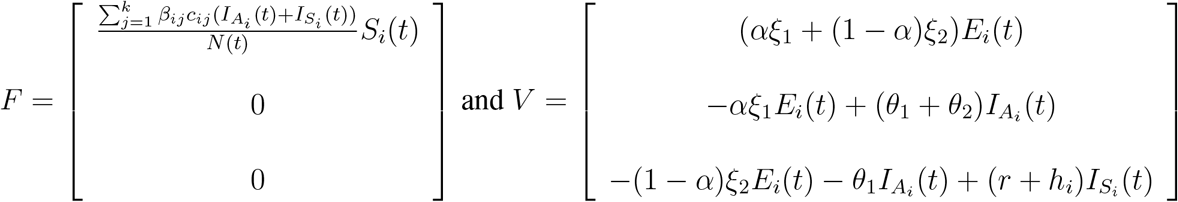

In order to compute 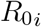 let the gradient of F be define as 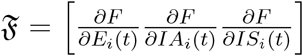 and let the gradient of V be define as 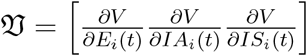 then we get:

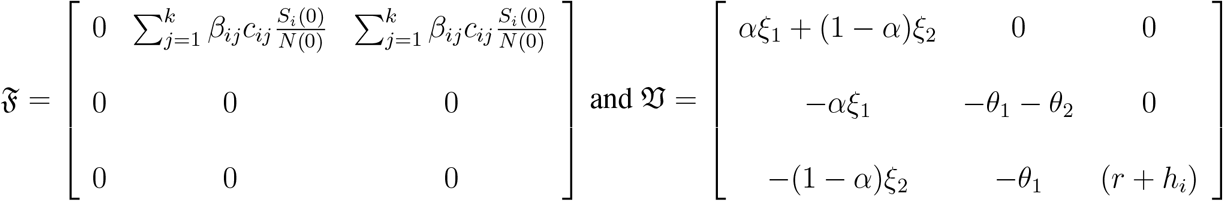

Then 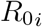 is the spectral radius of the second generation operator *ρ*(𝔉 𝔙^−1^) also know as the dominant eigenvalue of the matrix 𝔉 𝔙^−1^ Hence,

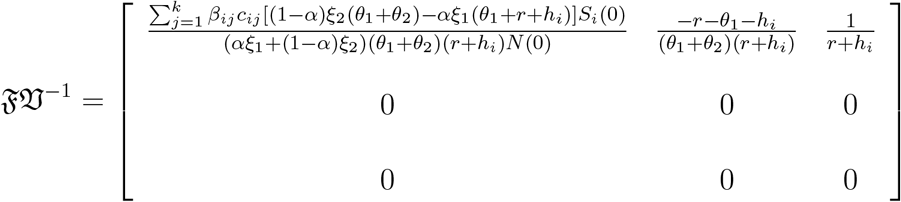

Then the dominant eigenvalue of 𝔉 𝔙^−1^ defined as *ρ*(𝔉 𝔙^−1^) is the 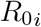 which means that the basic reproductive number for each age group *i* is:

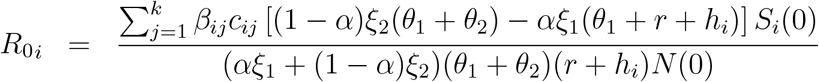

## C Comparison of different closure strategies (1 week, 2 weeks, 3 weeks, 4 weeks, 8 weeks and 24 weeks of social distancing)

## References

Anderson, Roy M, & May, RM. 1991. Infectious diseases of humans. 1991. New York: Oxford Science Publication Google Scholar.

Araz, Ozgur M., Damien, Paul, Paltiel David A., Burke, Sean, van de Geijn, Bryce, Galvani, Alison, & Meyers Lauren Ancel. 2012. Simulating school closure policies for cost effective pandemic decision making. BMC Public Health, 12(1), 449.

Araz, Ozgur M, Choi, Tsan-Ming, Olson David L, & Salman, F Sibel. 2020. Role of Analytics for Operational Risk Management in the Era of Big Data. Decision Sciences.

CDC, Centers for Disease Control and Prevention. 2020a. CDC COVID Data Tracker. https://www.cdc.gov/covid-data-tracker/cases. Date retrieved: June, 2020.

CDC, Centers for Disease Control and Prevention. 2020b. Coronavirus disease 2019 (COVID-19) how COVID-19 spreads. https://www.cdc.gov/coronavirus/2019-ncov/about/transmission.html. Date retrieved: March 13, 2020.

CDC, Centers for Disease Control and Prevention. 2020c. Coronavirus disease 2019 (COVID-19) situation sum-mary. https://www.cdc.gov/cor onavirus/2019-nCoV/summary.html. Date retrieved: March 13, 2020.

Ciavarella, C, Fumanelli, L, Merler, S, Cattuto, C, & Ajelli, M. 2016. School closure policies at municipality level for mitigating influenza spread: a model-based evaluation. BMC Infect Dis, 16.

Courtemanche, Charles, Garuccio, Joseph, Le, Anh, Pinkston, Joshua, & Yelowitz, Aaron. 2020. Strong Social Distancing Measures In The United States Reduced The COVID-19 Growth Rate. Health Affairs, 39(7), 10.1377/hlthaff.2020.00608. PMID: 32407171.

Dimitrov, Nedialko, Goll, Sebastian, Meyers, Lauren Ancel, Pourbohloul, Babak, & Hupert, Nathaniel. 2009. Optimizing tactics for use of the US antiviral strategic national stockpile for pandemic (H1N1) Influenza, 2009. PLoS currents, 1.

Duijzer, Lotty E, van Jaarsveld, Willem L, Wallinga, Jacco, & Dekker, Rommert. 2018a. Dose-optimal vaccine allocation over multiple populations. Production and Operations Management, 27(1), 143–159.

Duijzer, Lotty Evertje, van Jaarsveld, Willem, & Dekker, Rommert. 2018b. The benefits of combining early aspecific vaccination with later specific vaccination. European Journal of Operational Research, 271(2), 606–619.

Fumanelli, Laura, Ajelli, Marco, Merler, Stefano, Ferguson, Neil M., & Cauchemez, Simon. 2016. Model-Based Comprehensive Analysis of School Closure Policies for Mitigating Influenza Epidemics and Pandemics. PLOS Computational Biology, 12(1), 1–15.

Gojovic, MZ, Sander, B, Fisman, D, Krahn, MD, & CT, Bauch. 2009. Modelling mitigation strategies for pandemic (H1N1) 2009. CMAJ, 181(10), 673–680.

Griffiths, Jeff, Lowrie, Dawn, & Williams, Janet. 2000. An age-structured model for the AIDS epidemic. European Journal of Operational Research, 124(1), 1–14.

Lauer, Stephen A, Grantz Kyra H, Bi, Qifang, Jones Forrest K, Zheng, Qulu, Meredith Hannah R, Azman Andrew S, Reich Nicholas G, & Lessler, Justin. 2020. The incubation period of coronavirus disease 2019 (COVID-19) from publicly reported confirmed cases: estimation and application. Annals of internal medicine, 172(9), 577–582.

Lee, BY, Brown, ST, Cooley, P, & et al. 2010. Simulating school closure strategies to mitigate an influenza epidemic. J Public Health Manag Pract, 16(3), 252–261.

Li, Qun, Guan, Xuhua, Wu, Peng, Wang, Xiaoye, Zhou, Lei, Tong, Yeqing, Ren, Ruiqi, Leung Kathy SM, Lau Eric HY, Wong Jessica Y, et al. 2020a. Early transmission dynamics in Wuhan, China, of novel coronavirus– infected pneumonia. New England Journal of Medicine.

Li, Ruiyun, Chen, Bin, Song, Yimeng, Zhang, Tao, & Shaman, Jeffrey. 2020b. Substantial undocumented infection facilitates the rapid dissemination of novel coronavirus (SARS-CoV2). Science.

Linton, N.M., Kobayashi, T., Yang, Y., Hayashi, K., Akhmetzhanov, A.R., Jung, S.M., Yuan, B., Kinoshita, R., & Nishiura, H. 2020. Incubation Period and Other Epidemiological Characteristics of 2019 Novel Coronavirus Infections with Right Truncation: A Statistical Analysis of Publicly Available Case Data. Journal of Clinical Medicine, 9(538).

Mizumoto, Kenji, Kagaya, Katsushi, Zarebski, Alexander, & Chowell, Gerardo. 2020. Estimating the asymptomatic proportion of coronavirus disease 2019 (COVID-19) cases on board the Diamond Princess cruise ship, Yokohama, Japan, 2020. Eurosurveillance, 25(10), 2000180.

Mossong Joël, Hens, Niel, Jit, Mark, Beutels, Philippe, Auranen, Kari, Mikolajczyk, Rafael, Massari, Marco, Salmaso, Stefania, Tomba, Gianpaolo Scalia, Wallinga, Jacco, et al. 2008. Social contacts and mixing patterns relevant to the spread of infectious diseases. PLoS medicine, 5(3).

Ramirez-Nafarrate, Adrian, Araz Ozgur M, & Fowler John W. 2019. Decision assessment algorithms for location and capacity optimization under resource shortages. Decision Sciences.

Sander, Beate, Kwong Jeffrey C., Bauch Chris T., Maetzel, Andreas, McGeer, Allison, Raboud Janet M., & Krahn, Murray. 2010. Economic Appraisal of Ontario’s Universal Influenza Immunization Program: A Cost-Utility Analysis. PLOS Medicine, 7(4), 1–11.

Teytelman, Anna, & Larson Richard C. 2012. Modeling influenza progression within a continuous-attribute heterogeneous population. European journal of operational research, 220(1), 238–250.

Van den Driessche, Pauline, & Watmough, James. 2002. Reproduction numbers and sub-threshold endemic equilibria for compartmental models of disease transmission. Mathematical Biosciences, 180(1-2), 29–48.

Verity, R., Okell, L.C., Dorigatti, I., Winskill, P., Whittaker, C., Imai, N., Cuomo-Dannenburg, G., Thompson, H., Walker, P.G.T., Fu, H., Dighe, A., Griffin, J.T., Baguelin, M., Bhatia, S., Boonyasiri, A., Cori, A., Cucunuba, Z., FitzJohn, R., Gaythorpe, K., Green, W., Hamlet, A., Hinsley, W., Laydon, D., Nedjati-Gilani, G., Riley, S., van Elsland, S., Volz, E., Wang, H., Wang, Y., Xi, X., Donnelly, C.A., Ghani, A.C., & Ferguson, N.M. 2020. Estimates of the severity of coronavirus disease 2019: a model-based analysis. Lancet Infect Dis, 20(6), 669–677.

